# Effect of Lowering the Drink-Driving Blood Alcohol Limit in Scotland on Road Traffic Crashes: a Synthetic Difference-in-Differences Study

**DOI:** 10.64898/2026.06.18.26355950

**Authors:** Melina Jafari, Anupriya, Daniel J. Graham

**Affiliations:** Transport Strategy Centre, Imperial College London, London, SW7 2AZ, UK

## Abstract

**Objective:** To evaluate the road safety impact arising from Scotland’s 2014 reduction in the legal blood alcohol concentration (BAC) limit for drivers, and to assess whether the effect of the reform varied across different spatial contexts.

**Design:** A quasi-experimental statistical longitudinal study using a Synthetic Difference-in-Differences (SDID) approach.

**Setting:** Small-area panel data for Great Britain, with areas (Middle-layer Super Output Areas, MSOAs, in England and Wales and Intermediate Zones, IZs, in Scotland) classed into control and treatment groups according to whether they were exposed to Scotland’s BAC reform. The control and treatment groups comprise 7088 spatial units in England and Wales and 852 spatial units in Scotland, respectively, observed over the period 2008– 2019.

**Participants:** The study primarily analyses police-reported road traffic collision data from the UK Department for Transport’s STATS19 system. Data were analysed at the MSOA/ IZ level. This is a secondary dataset, and we therefore did not involve patients or the public in formulating the research question, determining outcome measures, or designing and conducting the study.

**Main Outcome Measures:** The main outcome measures were log-transformed rates of total road traffic crashes, and (weekend) night-time crashes (22:00–04:00) per 100,000 population. The latter is used as a proxy measure for drunk driving.

**Results:** Our results indicate that the reduction in the legal BAC limit led to statistically significant declines in road traffic crash rates. Aggregate estimates suggest reductions of 12.0% (95% confidence interval (CI): [-13.7%, -10.3%]) in total crashes, 15.6% (95% CI: [-20.7%, -10.2%]) in night-time crashes, and 12.4% (95% CI: [-16.7%, -7.9%]) in weekend night-time crashes. We also find substantial heterogeneity in treatment effects across spatial contexts. Effects were strongest in rural and less densely populated areas, where reductions exceeded 16% (95% CI: [-18.7%, -13.9%]) for total crashes and reached up to 29.6% (95% CI: [-35.8%, -22.8%]) for night-time and 21.4% (95% CI: [-28.3%, -13.9%]) for weekend night-time crashes. Moderate but statistically significant effects were also observed in dense urban areas, whereas effects in suburban and transitional areas were smaller and not statistically significant.

**Conclusions:** Our analysis suggests that lowering the legal BAC limit in Scotland led to meaningful reductions in road traffic crashes, particularly during higher-risk periods and in rural areas. The findings further suggest that the effectiveness of BAC regulation may vary across local contexts, highlighting the importance of accounting for spatial heterogeneity when evaluating road safety policies.

WHAT IS ALREADY KNOWN ON THIS TOPIC

- Lower legal blood alcohol concentration (BAC) limits for drivers have generally resulted in reductions in alcohol-related road traffic crashes in many settings.
- Scotland’s 2014 reduction in the legal BAC limit is a recent and policy-salient reform within Great Britain, making it an important case for assessing the real-world road safety effects of lower BAC thresholds.
- Previous evaluations of Scotland’s reform have reported limited or no statistically significant effects on road traffic crashes, but these studies have generally relied on relatively short post-intervention periods and data aggregated to larger spatial units, which may obscure variation in policy effects across areas.

WHAT THIS STUDY ADDS

- Using small-area panel data and a robust quasi-experimental statistical modelling approach, this study provides new evidence on the direct marginal effect of Scotland’s 2014 BAC reform on road traffic crashes.
- The reform is found to have led to significant reductions in total, night-time, and weekend night-time crash rates, with particularly strong effects in rural and less densely populated areas.
- These findings suggest that the effects of BAC policy are spatially heterogeneous and that analyses using shorter follow-up periods or more highly aggregated spatial data may underestimate, or fail to detect, important reductions in crash risk.

## 1 Introduction

Road traffic collisions represent one of the most serious and persistent global public health concerns. According to the World Health Organisation (WHO), approximately 1.19 million people die each year as a result of road traffic collisions, while an additional 20–50 million people suffer non-fatal injuries, many of which lead to long-term disability [1]. Beyond the human toll, these injuries impose a substantial economic cost on societies, with global losses estimated at around 3% of the gross domestic product (GDP) in most countries due to medical costs, lost productivity, and the wider economic consequences of deaths and disabilities [1]. Among the behavioural factors contributing to this burden, driving under the influence of alcohol remains one of the most important and preventable. In recognition of this, legal blood alcohol concentration (BAC) limits have become a central policy instrument in road safety regulation, although the thresholds applied vary widely across countries and regions. Following Scotland’s reduction of the legal BAC limit for drivers from 0.08 to 0.05 g/dL in December 2014 [2], doctors at the British Medical Association in 2023 called for the legal drink-driving limit in England, Wales, and Northern Ireland to be lowered to align with Scotland and with much of the rest of Europe [3]. This renewed debate is particularly crucial because, apart from Scotland, most of Great Britain continues to operate with one of the highest legal BAC limits in Europe. More broadly, cross-country variation in BAC thresholds remains substantial, with many jurisdictions adopting limits of 0.05 g/dL or lower, and some operating near-zero or zero-tolerance policies (Figure 1) [4]. Against this backdrop, understanding whether lower legal BAC limits translate into measurable improvements in road safety remains a pressing public health and policy question.

**Figure 1:**
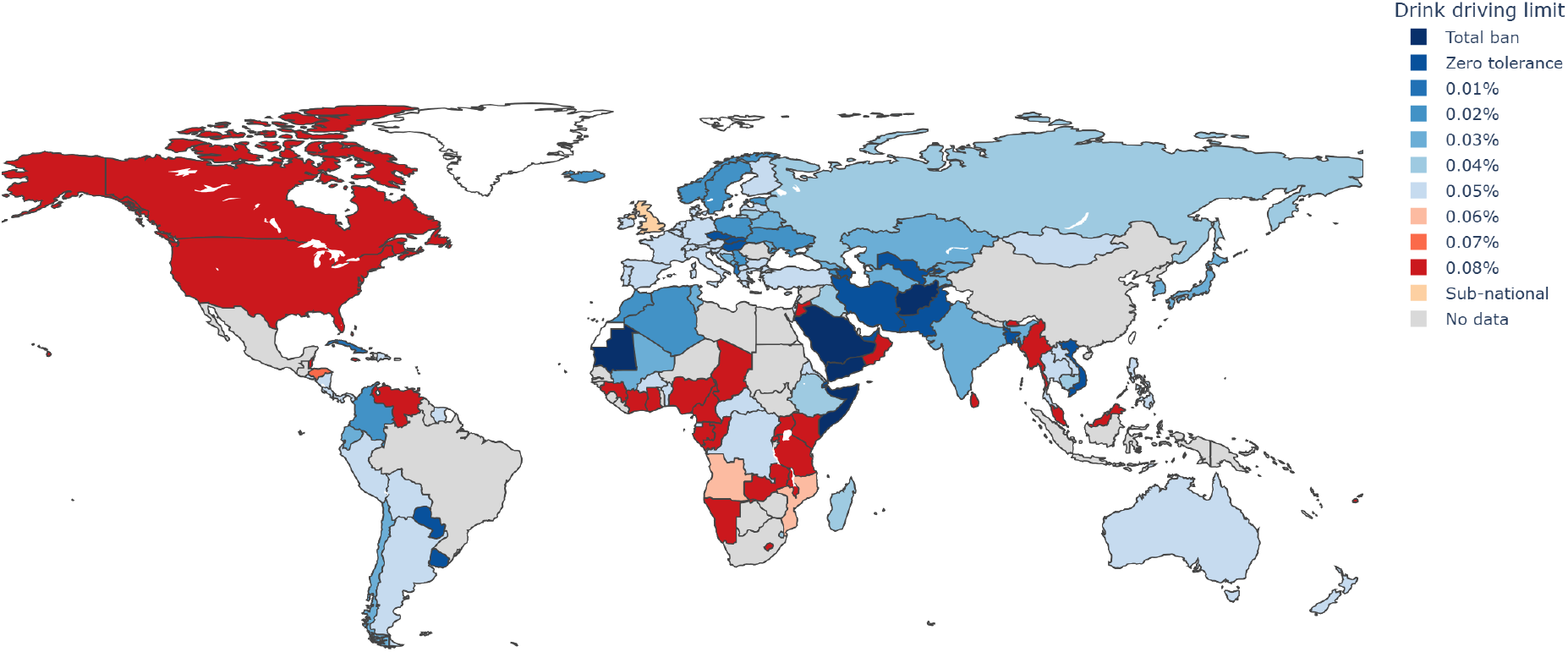
Drink Driving Limit by Country (2019), Source: World Health Organisation (WHO).

The rationale for regulating drink-driving through BAC thresholds is grounded in a large body of biological, behavioural, and epidemiological evidence. Alcohol impairs reaction time, coordination, attention, and judgment, while increasing risk-taking behaviour and reducing alertness, all of which undermine the ability to drive safely [5, 6, 7, 8]. BAC, typically measured through blood, urine, or breath samples, is the standard metric used to quantify alcohol exposure for legal and clinical purposes [9]. Evidence suggests that crash risk begins to increase even at relatively low BAC levels and rises sharply thereafter, often in a non-linear or exponential manner [10, 11, 12, 13]. The WHO has further estimated that up to 35% of worldwide daily road traffic fatalities are alcohol-related [14]. Recent work also suggests that alcohol-related crashes, while representing a minority of total crashes, are disproportionately severe and more likely to result in fatal outcomes, particularly at night and in rural areas [15]. Meta-analytic evidence similarly supports a strong and increasing relationship between alcohol consumption and crash risk, with even low levels of drinking linked to elevated injury risk and substantially higher risks at moderate and high levels of consumption [16]. Taken together, this literature provides a strong substantive basis for expecting that lower legal BAC thresholds may improve road safety.

Consistent with this expectation, substantial international literature has examined whether reducing legal BAC limits leads to fewer alcohol-related crashes and fatalities. Evidence from the United States suggests that lowering the legal BAC limit to 0.08 g/dL has led to reductions in fatal crashes typically ranging from 5% to 16% [17, 18, 19, 20]. Strong evidence has also emerged from Australia, where the adoption of a 0.05 g/dL limit, particularly alongside enforcement measures such as Random Breath Testing, has been linked to substantial reductions in alcohol-related crashes [21, 22]. Similar patterns have been reported in France, Japan, and other countries across Europe and Asia, where BAC reductions have been linked to declines in alcohol-related collisions and fatalities [23, 24, 25, 26]. However, despite this broadly supportive evidence, the magnitude of estimated effects varies considerably across studies and contexts. Some evaluations report negligible or null effects, including studies from Denmark, Great Britain, and the United States [27, 28, 29, 30, 31]. Overall, meta-analyses conclude that reducing legal BAC limits generally yields a reduction in alcohol-related crashes, often in the range of 5–11%, especially when legislative change is accompanied by visible enforcement and public awareness [32, 33]. Yet these reviews also emphasise that effect estimates are highly sensitive to enforcement intensity, behavioural adaptation, concurrent road-safety interventions, and methodological choices [10, 20, 32, 33].

This variation in findings reflects not only genuine differences across institutional and behavioural contexts, but also persistent methodological challenges in the literature. Much of the evidence on alcohol-related crash risk and BAC thresholds is derived from observational and regression-based approaches, including logistic regression, panel models, case-control studies, and quasi-experimental exposure designs [15, 34, 35]. While these methods have provided valuable insights, they are vulnerable to omitted variable bias and unobserved heterogeneity, as factors such as enforcement intensity, road conditions, weather, local travel behaviour, and driver risk preferences are often difficult to measure comprehensively. Case-control and quasi-induced exposure studies frequently rely on strong assumptions about the representativeness of control groups and exposure patterns, creating scope for selection bias [12, 34, 11]. Experimental and simulation-based studies have also deepened understanding of the physiological and behavioural effects of alcohol on driving performance, but their external validity is limited because simulated settings cannot fully reproduce real-world traffic conditions, enforcement environments, and driver decision-making [36, 37, 38, 39]. More generally, differences in data sources, outcome definitions, model specification, aggregation level, and contextual conditions contribute to substantial variation in estimated effects across studies [16]. These concerns become especially important when the goal is not simply to document alcohol-related crash risk, but to estimate the causal effect of a change in legal BAC limits.

In this setting, the central empirical challenge is to determine how collision outcomes would have evolved in the absence of the policy change. A large share of the policy evaluation literature relies on before–after comparisons or Difference-in-Differences (DiD) type designs [26, 28, 29, 31]. The underlying idea is to compare changes in outcomes in a treated jurisdiction with changes observed in untreated comparison areas, using the latter to approximate the counterfactual trajectory that the treated area would have followed in the absence of reform. However, the credibility of this approach depends on strong assumptions, most notably that treated and control units would have followed similar outcome trends in the absence of treatment. In practice, this assumption may be difficult to sustain if treated and comparison areas differ systematically in travel behaviour, enforcement intensity, urban form, exposure to other road-safety policies, or longer-term collision trends. Studies based on aggregate data or simple before–after comparisons may therefore struggle to isolate the independent effect of BAC reforms from other coincident changes, while even more sophisticated econometric approaches may remain sensitive to specification choices and the construction of the comparison group [10, 20, 32, 33]. For this reason, the existing evidence on BAC policy effectiveness, though substantial, remains less definitive than the broader biological and epidemiological case for stricter drink-driving regulation.

Scotland provides a particularly important setting in which to revisit this question. In December 2014, Scotland reduced the legal BAC limit for drivers from 0.08 to 0.05 g/dL of blood, while England and Wales retained the higher limit [2]. This reform created a highly relevant policy contrast within Great Britain and has since been repeatedly invoked in debates over whether the rest of the UK should follow suit [3]. Two prominent studies have examined the Scottish reform, both reporting limited or no statistically significant effects. Haghpanahan et al. [29], using a comparative interrupted time-series design with weekly country-level data from 2013 to 2016, find no evidence of a reduction in road traffic collisions following the policy change. Francesconi and James [28], using monthly local-authority-level data from 2009 to 2016, similarly reported no significant impact and suggested that weak enforcement and limited behavioural responses may have constrained the policy’s effectiveness. These findings are important, but they do not necessarily imply that lower BAC limits are ineffective in the Scottish context. First, both studies rely on relatively short post-treatment periods, which may be insufficient to detect longer-term behavioural adjustments or delayed policy effects. Second, analyses at the country or local-authority level may mask substantial heterogeneity in traffic conditions, alcohol use, enforcement visibility, and behavioural responses across smaller areas. Third, the DiD-based identification strategies used in prior work rely on assumptions about untreated trends that may be difficult to justify if treated and control regions differ systematically. Finally, one prior study classifies alcohol-related crashes using breath-test outcomes recorded in police crash data, where a crash is identified as drink-driving only if a driver fails or refuses a breath test [28]. Because testing and reporting may be incomplete, this outcome may understate the true incidence of alcohol involvement and potentially bias estimated policy effects.

In light of these limitations, there remains a need for evaluations based on longer time horizons, more spatially disaggregated data, and causal inference methods that improve the construction of the counterfactual. This study addresses these gaps by examining the impact of Scotland’s 2014 BAC reduction using a panel dataset derived from the *UK Department for Transport’s (DfT) STATS19 system*, aggregated at the Middle-layer Super Output Area (MSOA) level for England and Wales and the Intermediate Zone (IZ) level for Scotland over the period 2008–2019. The primary outcome is the logarithm of total crashes per 100,000 population. Secondary outcomes include the logarithm of night-time crashes and weekend night-time crashes per 100,000 population.

To estimate the causal effect of the reform, we apply the Synthetic Difference-in-Differences (SDID) method, which improves on conventional DiD by assigning weights to control units so as to better match pre-treatment outcome trajectories and thereby enhance the credibility of the counterfactual. In addition to estimating average treatment effects, we examine heterogeneous treatment effects by grouping spatial units with similar characteristics using a hierarchical clustering method and then re-estimating the SDID model within each cluster. This allows us to assess whether the policy had differential impacts across distinct types of areas, rather than assuming a uniform effect across space. Our primary objective is therefore to estimate whether Scotland’s BAC reform had a causal effect on road traffic collision outcomes, and our secondary objective is to assess whether any such effect varied systematically across different area types. By doing so, the study seeks to provide more robust and policy-relevant evidence for an ongoing public health debate in the United Kingdom.

## 2 Methods

This study adopts a quasi-experimental longitudinal panel design, exploiting Scotland’s 2014 reduction in the legal blood alcohol concentration (BAC) limit as a natural experiment. The analysis uses routinely collected police-reported collision data from Great Britain between 2008 and 2019 and applies a Synthetic Difference-in-Differences (SDID) framework to estimate the causal impact of the intervention on road safety. Secondary analyses assess whether the estimated effects vary across different area types.

### 2.1 Data Sources and Study Sample

The primary data source is the UK Department for Transport’s STATS19 system, which records police-reported road traffic collisions in Great Britain.^1^ We use collision records from 2008 to 2019. Each collision is assigned to a small-area spatial unit using its recorded geographic coordinates and fixed 2011 census geographies: Middle-layer Super Output Areas (MSOAs) for England and Wales^2^ and Intermediate Zones (IZs) for Scotland.^3^ Using fixed 2011 boundaries ensures temporal consistency and avoids changes arising from later administrative boundary revisions.

Records with missing geographic coordinates are excluded. We also exclude records that cannot be matched to an MSOA or IZ following the spatial join. The resulting dataset covers 6,791 MSOAs in England, 410 MSOAs in Wales, and 1,268 IZs in Scotland. We then construct an annual area-level panel and restrict the analysis to spatial units with complete observations over the full study period, yielding a balanced panel of 852 treated units in Scotland and 7,088 control units in England and Wales. We use annual rather than monthly or quarterly data because annual aggregation improves comparability with the existing policy-evaluation literature and reduces noise arising from seasonal variation, short-run anomalies, and reporting inconsistencies in higher-frequency crash data.

The study period ends in 2019 to avoid confounding from the COVID-19 pandemic. Pandemic-related changes in travel demand, traffic volumes, mobility restrictions, and enforcement practices introduce major structural breaks unrelated to the BAC reform and would complicate causal interpretation.

In addition to collision records, we compile area-level variables describing demographic, socioeconomic, spatial, and transport-network characteristics that may be relevant to road traffic crash outcomes. Annual population estimates are obtained from the Office for National Statistics for MSOAs^4^ and from the National Records of Scotland for IZs.^5^ Local economic activity is measured using Gross Value Added (GVA).^6^ Socioeconomic disadvantage is captured using the Index of Multiple Deprivation (IMD).^7^ Area size is derived from 2011 boundary geometries, and settlement structure is characterised using a harmonised urban–rural classification based on the proportion of urban land within each spatial unit.^8^ ^9^

We also construct a measure of spatial accessibility and density, mean effective density (MED), as in [40], which combines population concentration within an area with accessibility to surrounding populations:

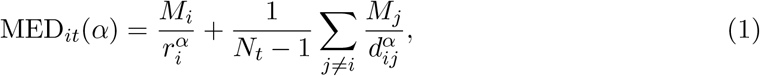

where *M*_*i*_ is the population of area *i, r*_*i*_ is its effective radius, and *d*_*ij*_ is the distance between areas *i* and *j*. This measure is intended to capture the geographic centrality of spatial units, their size distribution, and the surrounding distribution of economic mass.

Road-network characteristics are derived from the Ordnance Survey Open Roads dataset^10^ and aggregated to the MSOA/IZ level. These include total road length, segment count, road density, total nodes, mean node degree, true intersections, and intersection density. Population, GVA, IMD, and MED vary annually. Area size, urban–rural class, and road-network characteristics are treated as fixed structural characteristics over the study period. No missing observations remain in the final analytic sample.

We do not include covariates related to drink-driving enforcement, such as the number of patrol officers, drink-driving arrests, or alcohol testing rates. These factors may themselves respond to the policy change and are therefore better viewed as potential mediators or colliders rather than pre-treatment confounders. Conditioning on them could introduce collider bias and obscure part of the policy effect.

### 2.2 Treatment and Outcome Definition

The intervention being studied is Scotland’s reduction in the legal BAC limit for drivers from 0.08 to 0.05 g/dL, introduced in December 2014. Spatial units in Scotland define the treated group, while spatial units in England and Wales serve as controls because no equivalent change in BAC legislation occurs there during the study period. The main specification defines 2014– 2019 as the post-intervention period and 2008–2013 as the pre-intervention period. This coding reflects the possibility that behavioural responses, public awareness, and enforcement-related changes begin around the announcement and roll-out of the reform rather than only after its formal implementation. A sensitivity analysis using 2015–2019 as the post-intervention period is reported in the Appendix B.

We consider three related road safety outcomes: (i) the logarithm of total road traffic crashes per 100,000 population, (ii) the logarithm of night-time crashes per 100,000 population, and (iii) the logarithm of weekend night-time crashes per 100,000 population. The first outcome captures the overall road safety effect of the reform, while the latter two are designed to proxy alcohol-related crash risk.

Night-time crashes are defined as collisions occurring between 22:00 and 04:00. Weekend night-time crashes are defined as collisions occurring between 22:00 and 04:00 on Friday night/Saturday morning and between 22:00 and 04:00 on Saturday night/Sunday morning.

We use these temporal windows because directly recorded alcohol-related crashes are not publicly available in the STATS19 data, and because alcohol involvement in police-reported crash data is often incompletely observed even where such measures exist, owing to selective breath testing and reporting. In this context, time-based proxy outcomes provide a more consistent way to capture periods in which alcohol-related driving risk is likely to be elevated.

The choice of 22:00–04:00 is intended to capture the late-evening and early-morning period during which alcohol-related travel is more prevalent, while avoiding an overly narrow post-midnight definition that may miss relevant collisions occurring earlier in the night. The weekend night-time measure focuses more specifically on Friday and Saturday nights, when social drinking and associated road exposure are typically most concentrated. Similar temporal proxies are widely used in the literature on alcohol-related crash risk, although the precise definition of night-time varies across studies [41, 10, 12, 34, 18, 21, 22, 31, 42]. Our chosen windows therefore balance conceptual relevance, comparability with previous research, and the practical need for sufficiently stable small-area outcome counts.

We analyse log-transformed crash rates rather than raw counts. Because some small-area observations contain zero crashes, a constant of *ϵ* = 1 is added before log transformation. Population is used to standardise outcomes because consistent traffic-volume data are not available at the same spatial scale across Great Britain. The estimated effects are therefore interpreted as changes in population-standardised crash rates, recognising that population may not fully capture variation in traffic exposure.

### 2.3 Statistical Analysis

This study estimates the causal impact of Scotland’s BAC reform using the potential outcomes framework [43, 44, 45]. For each spatial unit *i* and year *t*, let *Y*_*it*_(1) denote the potential outcome that would be observed under the policy, and *Y*_*it*_(0) denote the potential outcome that would be observed in its absence. For each unit and year, only one of these two outcomes is observed. For treated units after the policy, *Y*_*it*_(1) is observed, whereas *Y*_*it*_(0) is unobserved and represents the counterfactual outcome. The central challenge of causal inference is therefore to estimate this missing counterfactual outcome.

The estimand of interest is the Average Treatment Effect on the Treated (ATT), which measures the average difference between the observed post-policy outcomes for treated units and the outcomes that would have occurred for those same units had the policy not been implemented. Formally, the ATT can be written as:

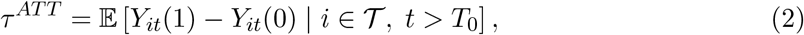

where *T* denotes the set of treated units and *T*_0_ denotes the final pre-treatment period. Since *Y*_*it*_(0) is not observed for treated units after the intervention, it must be estimated using information from untreated control units.

#### 2.3.1 Synthetic Difference-in-Differences

A standard Difference-in-Differences (DiD) approach estimates this counterfactual by comparing changes in outcomes over time between treated and control units [46, 47, 48, 49, 50, 51, 52, 53]. DiD is valid when, in the absence of treatment, treated and control units would have followed parallel trends. In practice, this assumption may be strong when treated and control areas differ systematically in their pre-policy trajectories. Synthetic Control Methods (SCM) address this concern by constructing a weighted combination of control units that closely reproduces the treated units’ pre-treatment outcome path [54, 55, 56, 57]. The synthetic control then serves as an estimate of the counterfactual trajectory for the treated group. However, conventional SCM is typically designed for settings with one or a small number of treated units and does not fully exploit the advantages of DiD-style time comparisons in panel settings with many treated and untreated units.

Synthetic Difference-in-Differences (SDID), proposed by Arkhangelsky et al. [58], combines the strengths of these two approaches. Like SCM, SDID assigns weights to control units so that the synthetic control group closely matches the treated group before the intervention. Like DiD, it compares treated and control outcomes before and after treatment and allows for additive unit and time effects. In addition, SDID introduces time weights, giving greater importance to pre-treatment periods that are most informative for the post-treatment counterfactual. Intuitively, the method first constructs a weighted comparison group from untreated areas in England and Wales that best reproduces the pre-policy outcome path of treated areas in Scotland, and then compares how treated and synthetic outcomes diverge after the reform.

Let *Y*_*it*_ denote the outcome for unit *i* = 1, …, *N* at time *t* = 1, …, *T* . Suppose that a subset of units *T* is exposed to treatment after time *T*_0_, while the remaining units *C* serve as controls. The SDID estimator can be expressed as a weighted difference-in-differences estimator:

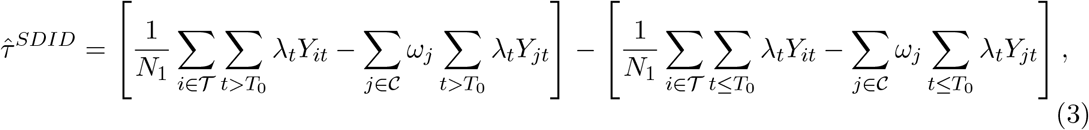

where *N*_1_ is the number of treated units, *ω*_*j*_ are unit weights assigned to control units, and *λ*_*t*_ are time weights assigned to pre-treatment periods. The unit weights construct a synthetic control group that resembles the treated group before the policy, while the time weights emphasise pre-treatment years that are most informative for predicting the post-treatment counterfactual. In this setting, the estimator measures whether crash outcomes in Scotland change after the BAC reform relative to a synthetic counterfactual constructed from comparable areas in England and Wales.

To further account for observed time-varying confounders, outcomes are residualised prior to SDID estimation. Specifically, for each outcome, an ordinary least squares regression is fitted using pre-treatment observations from control units only:

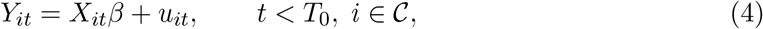

where *X*_*it*_ denotes the vector of observed covariates described in Section 2.1. Residualised outcomes are then defined as:

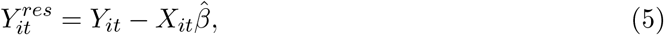

and SDID is applied to 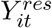 rather than the raw outcome. In the main specification, lagged values of the covariates are used in the residualisation step to preserve temporal ordering between covariates and outcomes and reduce the risk of post-treatment bias.

The validity of the SDID estimator relies on the assumption that, in the absence of the policy, treated and synthetic control units would have followed similar outcome trends. In this setting, this requires that no other major Scotland-specific policies affecting road safety are introduced contemporaneously with the BAC reform, and that spillover effects from Scotland to England and Wales are negligible. It also requires that the measurement of collision outcomes remains sufficiently stable over time and comparable across regions. By weighting control units to reproduce the pre-policy trajectory of the treated group, SDID is intended to strengthen the credibility of the counterfactual. Additional details on the choice of estimator, estimation of unit and time weights are provided in Appendix A.1 and Appendix A.2, respectively.

#### 2.3.2 Model fit and diagnostics

Model performance is evaluated using the root mean squared prediction error (RMSPE) in the pre-treatment period:

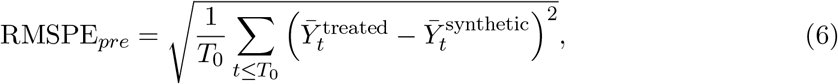

where 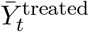 and 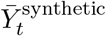 denote the average outcomes of treated units and their synthetic counterparts at time *t*. Lower values indicate better pre-intervention fit. To assess divergence following the intervention, we also calculate the ratio of post-treatment to pre-treatment RMSPE. Larger ratios indicate a stronger departure from pre-treatment trends after the policy implementation. In addition, graphical diagnostics are used to compare treated and synthetic control trends over time, providing a visual assessment of pre-treatment fit and post-treatment divergence.

There is no universally accepted numerical threshold for raw RMSPE in this context. Rather than relying on a fixed cutoff, RMSPE is used as a relative diagnostic of how well the synthetic control reproduces pre-treatment trends. This is because the magnitude of RMSPE depends on the scale and variability of the outcome, making absolute thresholds difficult to interpret across applications. Consistent with the literature following Abadie et al. [54] and Arkhangelsky et al. [58], emphasis is therefore placed on comparative and diagnostic evaluation, including the relative size of RMSPE, normalised fit measures, graphical inspection of trends, and placebo tests, rather than on strict numerical thresholds.

Because RMSPE is scale dependent, we also compute a normalised version of the pre-treatment RMSPE following Adhikari and Alm [59]. Specifically, the normalised RMSPE, or pre-treatment fit index, is defined as:

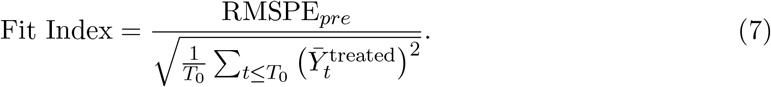

The denominator represents the benchmark RMSPE from a zero-fit model. This normalisation rescales RMSPE relative to the magnitude of the treated outcome series, allowing pre-treatment fit to be compared across outcomes with different scales and variances. A value of 0 indicates a perfect pre-treatment fit, values close to 0 indicate a strong fit, and values greater than 1 imply that the synthetic control performs worse than the benchmark zero-fit model. Although some studies have treated values below 0.10 as indicating strong pre-treatment fit [59], we do not use this as a strict acceptance criterion. Instead, the fit index is interpreted jointly with graphical diagnostics, placebo tests, and comparative RMSPE measures.

#### 2.3.3 Inference and implementation

Statistical significance of the estimated ATT is assessed using placebo, or permutation, tests, as described in Appendix A.3. Under the null hypothesis of no causal effect, assigning treatment randomly to units that were not actually exposed should generate estimated effects reflecting random variation rather than the BAC reform. By repeatedly generating such placebo assignments and re-estimating the model, we obtain an empirical distribution of treatment effects under the null. The observed SDID estimate is then compared with this placebo distribution.

In particular, we report a placebo-based standard error, calculated from the variability of the placebo estimates, and use this to construct an approximate 95% confidence interval around the observed SDID estimate (see Appendix A.3 for details). We also report a placebo (p)-value, defined as the proportion of placebo estimates that are at least as extreme as the observed estimate in the hypothesised direction of the policy effect. Because the policy is expected to reduce crash rates, inference is based on the lower tail of the placebo distribution: a small (p)-value indicates that few, if any, placebo assignments produce reductions as large as those observed for Scotland.

Finally, we report the mean of the placebo estimates, which summarises the average effect obtained under random reassignment, and the 95% placebo interval, which describes the range containing most placebo estimates. These quantities provide a benchmark for assessing whether the observed effect is unusually large relative to effects that could arise by chance.

All SDID models are implemented in Python using custom optimisation routines based on quadratic programming. Outcomes are not standardised prior to estimation, as scaling is handled implicitly through the weighting procedure. Unit and time weights are estimated via constrained minimisation with ridge regularisation, subject to non-negativity and adding-up constraints. The regularisation parameter *ζ* is calibrated based on the variability of pre-treatment outcomes following Arkhangelsky et al. [58].

Given the large number of control units, the donor pool is restricted using a prespecified pre-selection procedure based on similarity in pre-treatment trends. Specifically, control units are ranked according to their distance from the treated units in terms of pre-policy trajectories, and only those with the most similar trends are retained in the donor pool. This approach ensures that the synthetic control is constructed from units that provide a more credible counterfactual for the treated group. Models are then estimated iteratively using subsets of these pre-selected control units, and the preferred specification is chosen according to a prespecified model-selection rule that minimises pre-treatment RMSPE. Because this rule relies only on pre-treatment fit, it avoids selecting the model on the basis of post-treatment outcomes.

### 2.4 Heterogeneous Treatment Effects

Estimating only the average treatment effect may mask substantial variation in policy impacts across different types of areas. In the context of road safety interventions, the effectiveness of reductions in legal BAC limits is likely to depend on local characteristics such as population density, urban form, socioeconomic conditions, and road infrastructure. Urban areas with greater public transport availability and different travel patterns may respond differently to such policies than rural areas with higher car dependency and different enforcement contexts. Ignoring such heterogeneity may therefore obscure important differences in how the policy operates across space.

To examine heterogeneity in policy impacts, we estimate conditional average treatment effects on the treated (CATTs) across groups of areas with similar structural characteristics. Specifically, for each cluster *g*, the relevant estimand is:

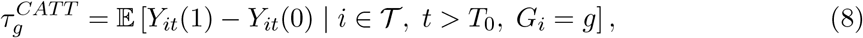

where *G*_*i*_ denotes the cluster membership of unit *i*. These CATTs measure whether the effect of Scotland’s BAC reform differs systematically across distinct area types.

To identify such area types, spatial units are grouped into clusters based on socioeconomic, demographic, and built-environment characteristics. All clustering variables, described in Section 2.1, are constructed using pre-treatment averages and are therefore treated as time-invariant area-level characteristics. This ensures that cluster assignment is not influenced by post-policy outcomes and avoids potential post-treatment bias.

Given the high dimensionality of these variables, Principal Component Analysis (PCA) is applied to reduce dimensionality while preserving most of the variation in the data [60]. The first seven principal components are retained, as they jointly explain at least 90% of the total variance in the clustering variables. Clustering is then performed using hierarchical agglomerative clustering with Ward’s linkage, which minimises within-cluster variance [61]. This approach is well-suited to the present setting for three reasons. First, the underlying number and shape of area types are not known a priori. Second, unlike partition-based methods such as k-means, hierarchical clustering is less sensitive to initial conditions and does not impose a strong assumption that clusters should be spherical or of similar size. Third, the hierarchical structure provides a transparent way to examine how observations group together across different levels of aggregation, which is useful when clustering areas defined by multiple spatial and socioeconomic characteristics. Although alternative clustering approaches are available, hierarchical clustering is preferred here because of its stability, interpretability, and flexibility in handling heterogeneous spatial data.

The optimal number of clusters is determined using two standard validation metrics: the silhouette score [62], with higher values indicating better-defined and more separated clusters, and the Davies–Bouldin index [63], with lower values indicating better clustering performance. To ensure reliable estimation, clusters are also required to contain a minimum number of treated and control units. Based on these criteria, the number of clusters is set to *k* = 5, yielding well-separated and internally coherent groups of spatial units.

The resulting clusters also exhibit substantial differences in the pre-treatment distributions and trajectories of the crash outcomes of interest. This is in line with the expectation that areas grouped on the basis of socioeconomic, demographic, and built-environment characteristics will face systematically different road safety conditions. The presence of such outcome heterogeneity further motivates the estimation of conditional ATTs across area types. Additional details on PCA, cluster profiles, and cluster-wise pre-treatment diagnostics are provided in Appendix A.4, Appendix A.6, and Appendix A.7, respectively.

Cluster labels are assigned based on the dominant socioeconomic and transport-network characteristics of each group. The resulting cluster definitions are summarised in Table 1. Furthermore, Figure 2 presents the spatial distribution of the resulting clusters across Great Britain, revealing substantial geographic heterogeneity between urban centres, semi-urban areas, and rural regions. Overall, the clustering identifies distinct area types that differ systematically in both socioeconomic conditions and transport-network characteristics. These differences provide a meaningful basis for examining heterogeneous responses to the BAC reform.

**Table 1:** Cluster definitions.

| Cluster | Label | Total Areas | Treated | Control |
| --- | --- | --- | --- | --- |
| 0 | Dense Urban Core | 5500 | 471 | 5029 |
| 1 | Large Rural Road-Network Areas | 321 | 78 | 243 |
| 2 | Suburban/Semi-Urban Areas | 225 | 6 | 219 |
| 3 | Urban–Rural Transition Areas | 75 | 21 | 54 |
| 4 | Typical Rural Areas | 1819 | 276 | 1543 |

**Figure 2:**
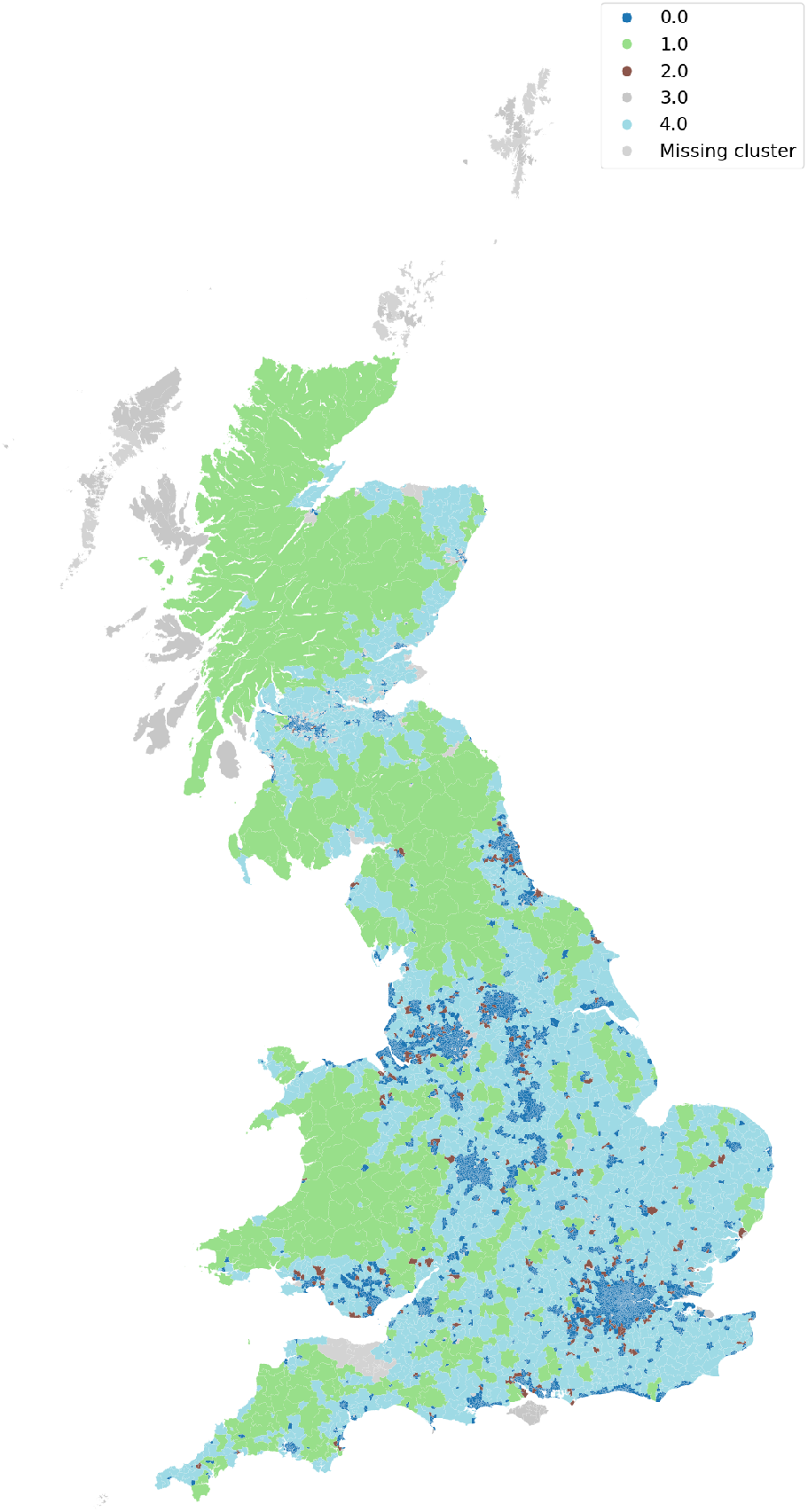
Spatial distribution of MSOA/IZ hierarchical clusters

Finally, the SDID model described in Section 2.3 is estimated separately within each cluster. For each cluster, treated units are compared with a weighted combination of control units belonging to the same cluster and therefore sharing similar structural characteristics. This restriction improves comparability and reduces bias arising from systematic differences across heterogeneous areas. The resulting cluster-specific estimates can therefore be interpreted as conditional ATTs for distinct types of areas.

### 2.5 Ethics Approval

Ethics approval is not required for this study because it uses publicly available, routinely collected, de-identified administrative data aggregated to area-year level and does not involve identifiable human participants.

### 2.6 Reporting Statement

This study is reported in accordance with the STROBE reporting guideline for observational studies. Given the use of routinely collected administrative data, relevant RECORD items are also addressed.

### 2.7 Patient and Public Involvement

No patients or members of the public are involved in setting the research question, determining outcome measures, or designing and conducting the study because the analysis uses secondary de-identified administrative data aggregated to area-year level.

## 3 Results

We first report the aggregate estimated effects of Scotland’s blood alcohol concentration (BAC) limit reduction policy on road traffic crash rates. We then examine whether these effects vary across clusters of spatial units defined by pre-treatment socioeconomic, demographic, and built-environment characteristics.

### 3.1 Aggregate Effects

Table 2 reports the aggregate estimated effects of Scotland’s BAC reduction policy across the full sample. Across all three outcomes, the policy is associated with lower crash rates after 2014. The estimated average treatment effect on the treated (ATT) corresponds to a 12.05% reduction in total crash rates, a 15.61% reduction in night-time crash rates, and a 12.38% reduction in weekend night-time crash rates. All three estimates are statistically significant under placebo-based inference, with placebo p-values of 0.0033.

**Table 2:** Aggregate treatment effects.

(a) Estimated ATT and model fit statistics
| Outcome | $\hat{\tau}^{SDID}$ | Pre-RMSPE | Fit Index | Post/Pre RMSPE Ratio | Policy Percent Effect |
| --- | --- | --- | --- | --- | --- |
| Log Total Crash Rate | -0.1284 | 0.0061 | 0.0593 | 24.2011 | -12.05% |
| Log Night Crash Rate | -0.1697 | 0.0119 | 0.0617 | 18.0668 | -15.61% |
| Log Weekend Crash Rate | -0.1322 | 0.0312 | 0.2623 | 4.8271 | -12.38% |

(b) Placebo-based inference
| Outcome | $\widehat{SE}$ | 95% CI based on Placebo SE | Placebo Mean | Placebo 95% Interval | Placebo p-value |
| --- | --- | --- | --- | --- | --- |
| Log Total Crash Rate | 0.0099 | [-0.1478, -0.1090] | 0.0132 | [-0.0071, 0.0308] | 0.0033 |
| Log Night Crash Rate | 0.0317 | [-0.2318, -0.1076] | 0.1670 | [0.1095, 0.2324] | 0.0033 |
| Log Weekend Crash Rate | 0.0255 | [-0.1822, -0.0822] | 0.0297 | [-0.0202, 0.0849] | 0.0033 |

Pre-treatment fit is strong for the aggregate models. Pre-intervention RMSPE values are low for all three outcomes, and the fit index indicates close agreement between treated units and their synthetic counterparts before the policy change for total and night-time crash rates. The fit index is somewhat larger for weekend night-time crashes, reflecting the greater volatility of this lower-frequency outcome, but the post/pre RMSPE ratio remains well above 1 for all three outcomes, indicating substantial post-treatment divergence relative to pre-treatment fit.

Placebo-based inference provides strong evidence that the observed reductions are unlikely to be generated by random assignment variation. The placebo means are positive for all three outcomes, suggesting that placebo-treated units did not exhibit comparable post-2014 reductions. In contrast, the observed SDID estimates are negative and lie well below the empirical placebo distributions. The one-sided lower-tail placebo *p*-values are equal to 0.0033 for all outcomes, indicating that none of the placebo assignments produced treatment effects as negative as the observed estimates. The placebo-based 95% confidence intervals are entirely below zero for all outcomes, further supporting a statistically significant crash-reducing effect of the BAC-limit reduction. In Appendix D.1 the placebo tau distribution is compared with the estimated treatment-effect distribution.

Figure 3 compares observed outcomes in treated units with their synthetic counterparts over time. The close alignment of the two series before 2014 indicates good pre-treatment fit. After the policy intervention, the treated series diverges from the synthetic control for all three outcomes, consistent with a policy-associated decline in crash rates.

**Figure 3:**
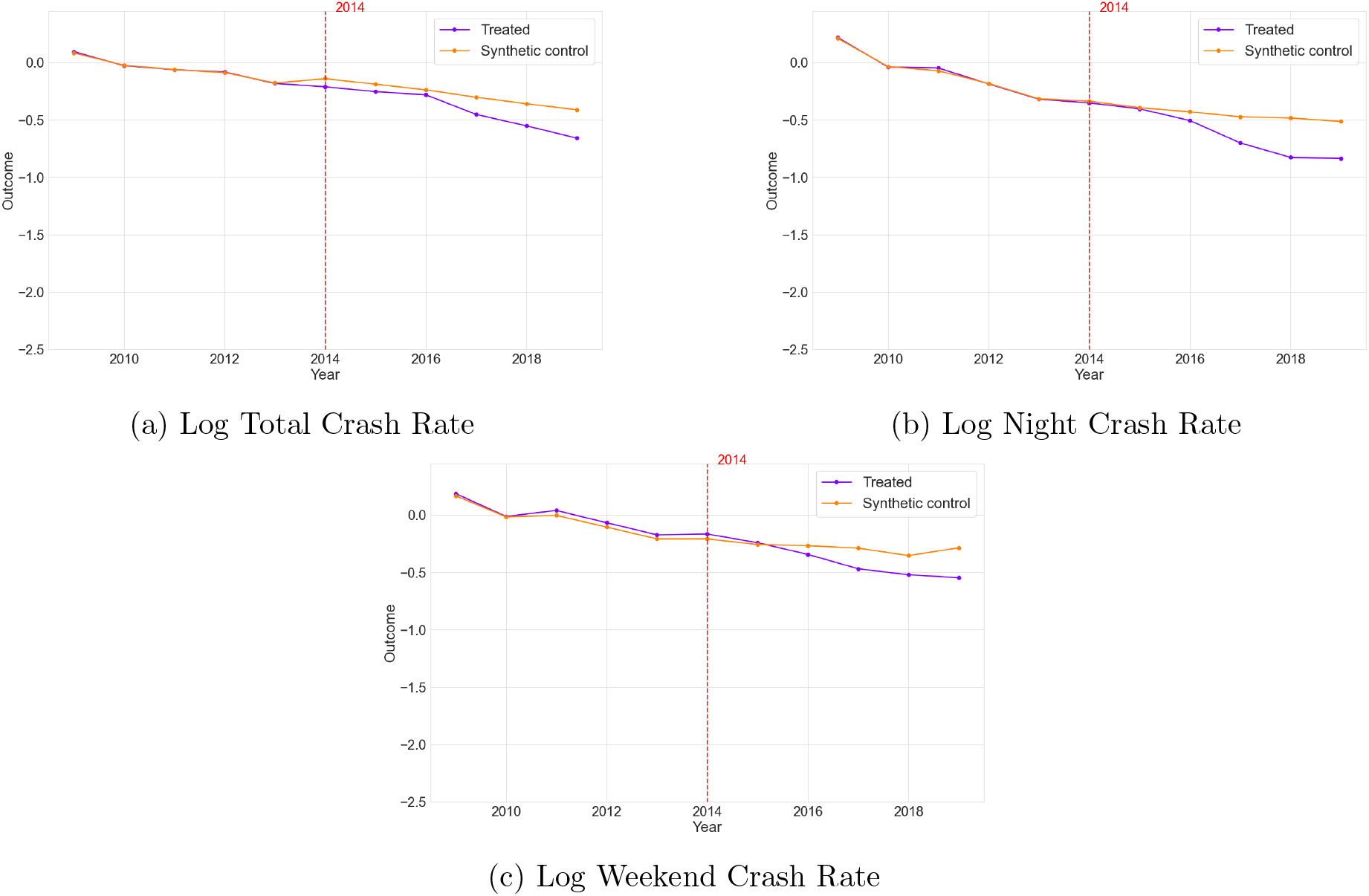
Observed and synthetic crash trends before and after BAC policy implementation: Aggregate effects

### 3.2 Heterogeneous Effects

The estimated effects of the BAC reduction policy vary substantially across area types. Table 3 reports cluster-specific conditional average treatment effects on the treated (CATTs). The largest and most precisely estimated reductions are observed in *Large Rural Road-Network Areas* (cluster 1) and *Typical Rural Areas* (cluster 4). In cluster 1, the estimated effects correspond to reductions of 21.03% in total crash rates, 47.61% in night-time crash rates, and 39.42% in weekend night-time crash rates. In cluster 4, the corresponding reductions are 16.33%, 29.62%, and 21.43%. All of these estimates are statistically significant under placebo-based inference and lie well below the empirical placebo distributions.

**Table 3:** Heterogeneous treatment effects.

(a) Estimated CATT and model fit statistics
| Cluster | Outcome | $\hat{\tau}^{SDID}$ | Pre-RMSPE | Fit Index | Post/Pre RMSPE Ratio | Policy Percent Effect |
| --- | --- | --- | --- | --- | --- | --- |
| 0 | Log Total Crash Rate | -0.0988 | 0.0072 | 0.0730 | 18.2206 | -9.40% |
|  | Log Night Crash Rate | -0.1427 | 0.0149 | 0.0777 | 12.3705 | -13.30% |
|  | Log Weekend Crash Rate | -0.0912 | 0.0269 | 0.2923 | 4.2105 | -8.71% |
| 1 | Log Total Crash Rate | -0.2360 | 0.0306 | 0.2325 | 8.3826 | -21.03% |
|  | Log Night Crash Rate | -0.6464 | 0.1336 | 0.4129 | 5.5812 | -47.61% |
|  | Log Weekend Crash Rate | -0.5012 | 0.0838 | 0.2391 | 6.4129 | -39.42% |
| 2 | Log Total Crash Rate | -0.0339 | 0.0377 | 0.1926 | 4.6727 | -3.33% |
|  | Log Night Crash Rate | -0.1784 | 0.1923 | 0.3342 | 3.9578 | -16.34% |
|  | Log Weekend Crash Rate | -0.2250 | 0.0752 | 0.1919 | 6.9043 | -20.15% |
| 3 | Log Total Crash Rate | -0.0558 | 0.1654 | 0.2942 | 1.4718 | -5.43% |
|  | Log Night Crash Rate | -0.2147 | 0.3354 | 0.7296 | 1.7164 | -19.32% |
|  | Log Weekend Crash Rate | 0.1194 | 0.2379 | 0.6326 | 1.0756 | 12.68% |
| 4 | Log Total Crash Rate | -0.1783 | 0.0178 | 0.0607 | 11.7237 | -16.33% |
|  | Log Night Crash Rate | -0.3513 | 0.0464 | 0.0936 | 8.9762 | -29.62% |
|  | Log Weekend Crash Rate | -0.2412 | 0.0302 | 0.0911 | 9.2470 | -21.43% |

(b) Placebo-based inference
| Cluster | Outcome | $\widehat{SE}$ | 95% CI based on Placebo SE | Placebo Mean | Placebo 95% Interval | Placebo p-value |
| --- | --- | --- | --- | --- | --- | --- |
| 0 | Log Total Crash Rate | 0.0130 | [-0.1243, -0.0733] | 0.0243 | [-0.0026, 0.0507] | 0.0033 |
|  | Log Night Crash Rate | 0.0398 | [-0.2207, -0.0647] | 0.1268 | [0.0478, 0.1998] | 0.0033 |
|  | Log Weekend Crash Rate | 0.0334 | [-0.1567, -0.0257] | 0.0561 | [-0.0138, 0.1228] | 0.0033 |
| 1 | Log Total Crash Rate | 0.0282 | [-0.2913, -0.1807] | -0.0004 | [-0.0552, 0.0521] | 0.0033 |
|  | Log Night Crash Rate | 0.1005 | [-0.8434, -0.4494] | 0.1102 | [-0.0958, 0.3001] | 0.0033 |
|  | Log Weekend Crash Rate | 0.1107 | [-0.7182, -0.2842] | 0.0100 | [-0.2167, 0.2186] | 0.0033 |
| 2 | Log Total Crash Rate | 0.1098 | [-0.2491, 0.1813] | 0.0112 | [-0.1953, 0.2164] | 0.3389 |
|  | Log Night Crash Rate | 0.3498 | [-0.8640, 0.5072] | 0.0268 | [-0.6714, 0.6523] | 0.2857 |
|  | Log Weekend Crash Rate | 0.3303 | [-0.8724, 0.4224] | -0.0137 | [-0.6993, 0.6522] | 0.2558 |
| 3 | Log Total Crash Rate | 0.0780 | [-0.2087, 0.0971] | -0.0097 | [-0.1663, 0.1332] | 0.2492 |
|  | Log Night Crash Rate | 0.2726 | [-0.7490, 0.3196] | -0.0005 | [-0.5292, 0.5087] | 0.2292 |
|  | Log Weekend Crash Rate | 0.2852 | [-0.4396, 0.6784] | 0.0217 | [-0.5919, 0.5688] | 0.6146 |
| 4 | Log Total Crash Rate | 0.0147 | [-0.2071, -0.1494] | 0.0168 | [-0.0114, 0.0469] | 0.0033 |
|  | Log Night Crash Rate | 0.0469 | [-0.4432, -0.2594] | 0.1088 | [0.0142, 0.1974] | 0.0033 |
|  | Log Weekend Crash Rate | 0.0469 | [-0.3331, -0.1493] | 0.0378 | [-0.0571, 0.1267] | 0.0033 |

**Table 4:**
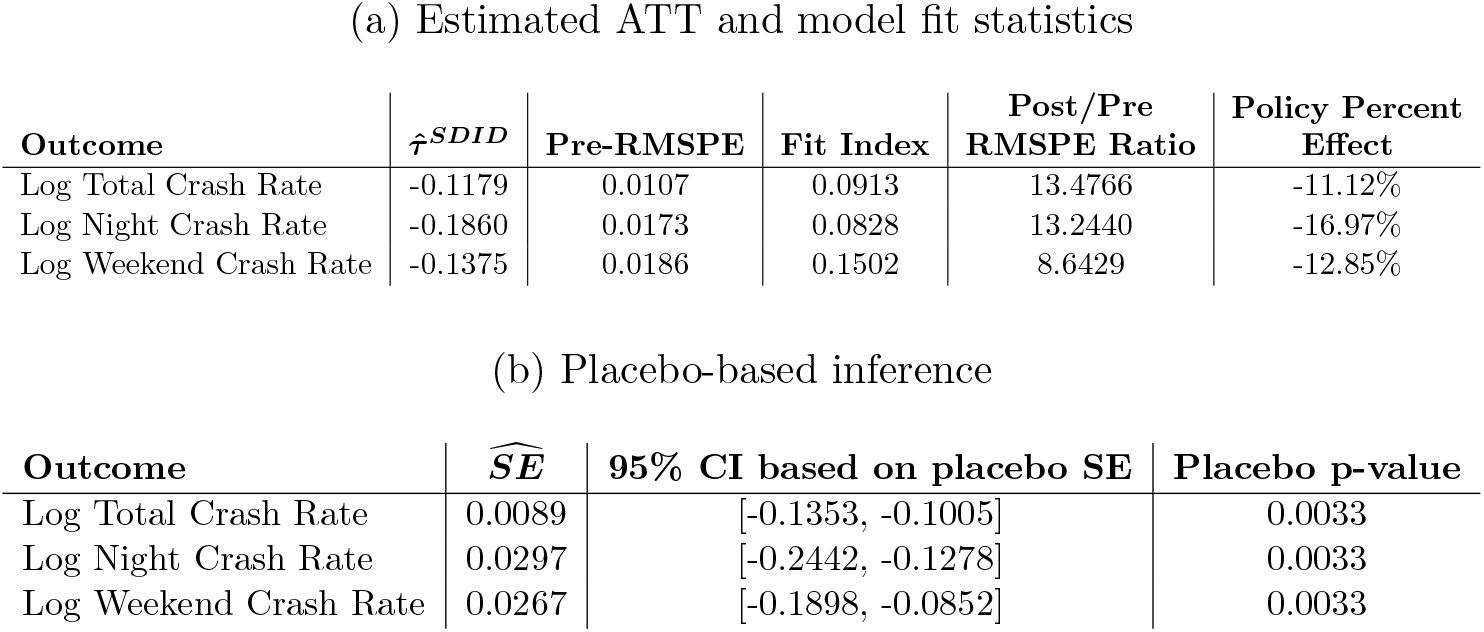
Aggregate Treatment Effects.

**Table 5:** Heterogeneous Treatment Effects.

(a) Estimated CATT and model fit statistics
| Cluster | Outcome | $\hat{\tau}^{SDID}$ | Pre-RMSPE | Fit Index | Post/Pre RMSPE Ratio | Policy Percent Effect |
| --- | --- | --- | --- | --- | --- | --- |
| 0 | Log Total Crash Rate | -0.0945 | 0.0079 | 0.0761 | 16.3995 | -9.01% |
|  | Log Night Crash Rate | -0.1558 | 0.0211 | 0.1023 | 9.5833 | -14.43% |
|  | Log Weekend Crash Rate | -0.0965 | 0.0275 | 0.2619 | 4.0182 | -9.20% |
| 1 | Log Total Crash Rate | -0.1827 | 0.0475 | 0.3033 | 4.5424 | -16.70% |
|  | Log Night Crash Rate | -0.6124 | 0.1288 | 0.3166 | 5.8280 | -45.79% |
|  | Log Weekend Crash Rate | -0.3949 | 0.0985 | 0.2780 | 4.6121 | -32.62% |
| 2 | Log Total Crash Rate | 0.0243 | 0.0393 | 0.1655 | 4.2847 | 2.46% |
|  | Log Night Crash Rate | -0.2279 | 0.1746 | 0.3399 | 4.6700 | -20.38% |
|  | Log Weekend Crash Rate | -0.1221 | 0.0941 | 0.2335 | 5.6623 | -11.50% |
| 3 | Log Total Crash Rate | -0.0654 | 0.1564 | 0.2757 | 1.7976 | -6.33% |
|  | Log Night Crash Rate | -0.3829 | 0.4108 | 0.8270 | 1.6748 | -31.81% |
|  | Log Weekend Crash Rate | 0.0578 | 0.2635 | 1.0477 | 0.8067 | 5.96% |
| 4 | Log Total Crash Rate | -0.1536 | 0.0340 | 0.1064 | 5.9263 | -14.24% |
|  | Log Night Crash Rate | -0.3762 | 0.0645 | 0.1267 | 6.7456 | -31.36% |
|  | Log Weekend Crash Rate | -0.3073 | 0.0588 | 0.1901 | 5.9227 | -26.45% |

(b) Placebo-based inference
| Cluster | Outcome | $\widehat{SE}$ | 95% CI based on placebo SE | Placebo p-value |
| --- | --- | --- | --- | --- |
| 0 | Log Total Crash Rate | 0.0129 | [-0.1198, -0.0692] | 0.0033 |
|  | Log Night Crash Rate | 0.0389 | [-0.2320, -0.0796] | 0.0033 |
|  | Log Weekend Crash Rate | 0.0344 | [-0.1639, -0.0291] | 0.0033 |
| 1 | Log Total Crash Rate | 0.0284 | [-0.2384, -0.1270] | 0.0033 |
|  | Log Night Crash Rate | 0.1046 | [-0.8174, -0.4074] | 0.0033 |
|  | Log Weekend Crash Rate | 0.1088 | [-0.6081, -0.1816] | 0.0033 |
| 2 | Log Total Crash Rate | 0.1067 | [-0.1848, 0.2334] | 0.8306 |
|  | Log Night Crash Rate | 0.3430 | [-0.9002, 0.4444] | 0.4917 |
|  | Log Weekend Crash Rate | 0.3496 | [-0.8073, 0.5631] | 0.6943 |
| 3 | Log Total Crash Rate | 0.0750 | [-0.2124, 0.0816] | 0.3787 |
|  | Log Night Crash Rate | 0.2973 | [-0.9656, 0.1998] | 0.2193 |
|  | Log Weekend Crash Rate | 0.2922 | [-0.5149, 0.6305] | 0.8438 |
| 4 | Log Total Crash Rate | 0.0141 | [-0.1812, -0.1260] | 0.0033 |
|  | Log Night Crash Rate | 0.0496 | [-0.4734, -0.2790] | 0.0033 |
|  | Log Weekend Crash Rate | 0.0506 | [-0.4065, -0.2081] | 0.0033 |

Moderate but statistically significant reductions are also observed in the *Dense Urban Core* cluster (cluster 0), where the estimated effects correspond to reductions of 9.40% in total crash rates, 13.30% in night-time crash rates, and 8.71% in weekend night-time crash rates. By contrast, the estimates for *Suburban/Semi-Urban Areas* (cluster 2) and *Urban–Rural Transition Areas* (cluster 3) are smaller and not statistically distinguishable from zero across all three outcomes, moreover, the placebo tau and the estimated treatment-effect distributions are overlapping. In cluster 3, the estimate for weekend night-time crashes is positive, but the confidence interval is wide and the placebo p-value indicates no statistically significant effect. Appendix D.2 compares the placebo tau distribution with the estimated treatment-effect distribution in each cluster.

Pre-treatment fit is strongest in clusters 0 and 4, where the fit indices are generally low across outcomes and the post/pre RMSPE ratios indicate clear post-treatment divergence. Fit is weaker in clusters 2 and 3, especially for night-time and weekend night-time crash outcomes, reflecting smaller treated samples and greater year-to-year volatility in these lower-frequency outcomes. These patterns are consistent with the wider confidence intervals and less precise inference observed in those clusters. Additional treated-versus-synthetic trend plots for each cluster are reported in Figures 4-8.

**Figure 4:**
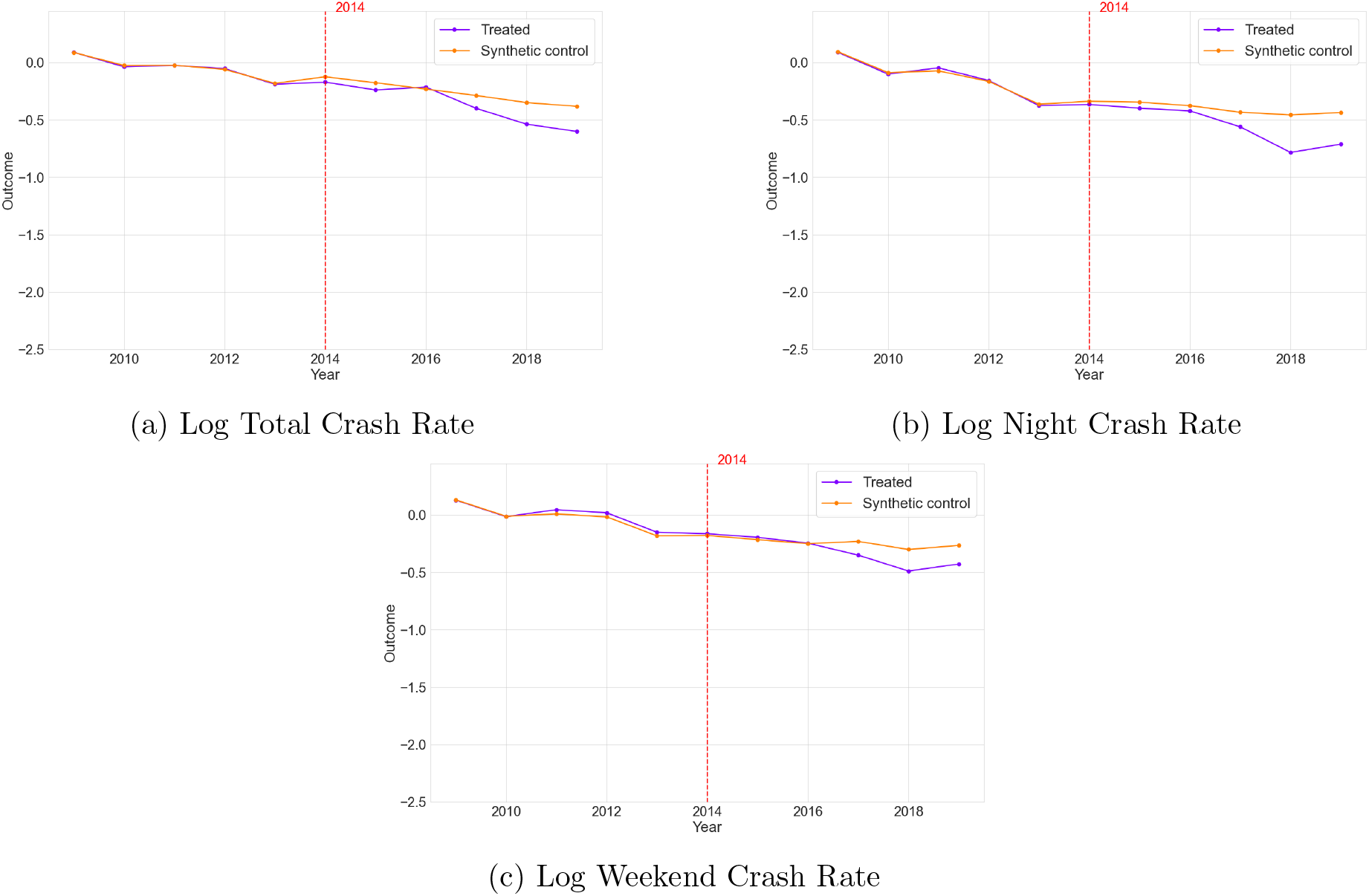
Observed and synthetic crash trends before and after BAC policy implementation: Heterogeneous effects, Cluster 0

**Figure 5:**
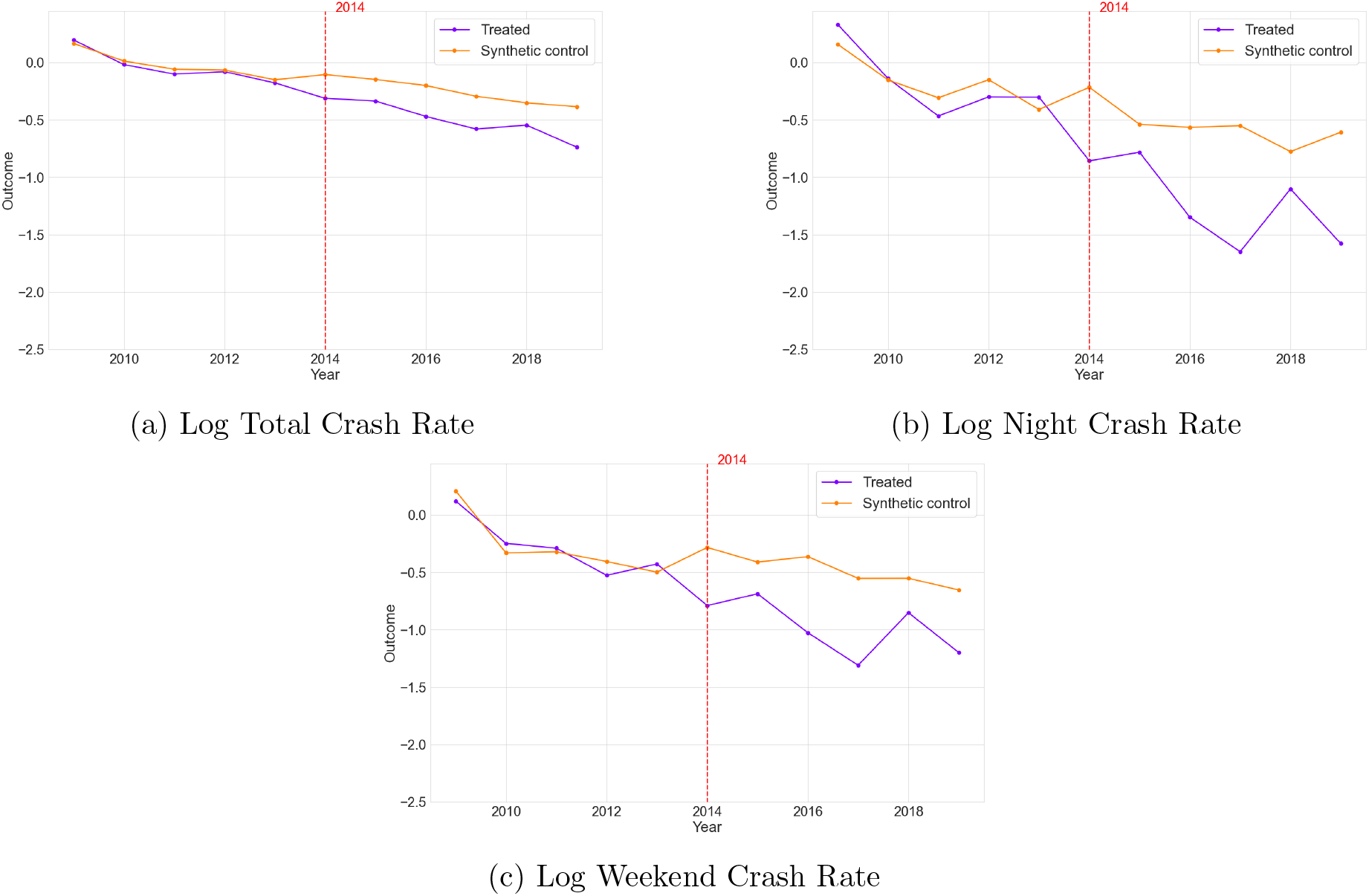
Observed and synthetic crash trends before and after BAC policy implementation: Heterogeneous effects, Cluster 1

**Figure 6:**
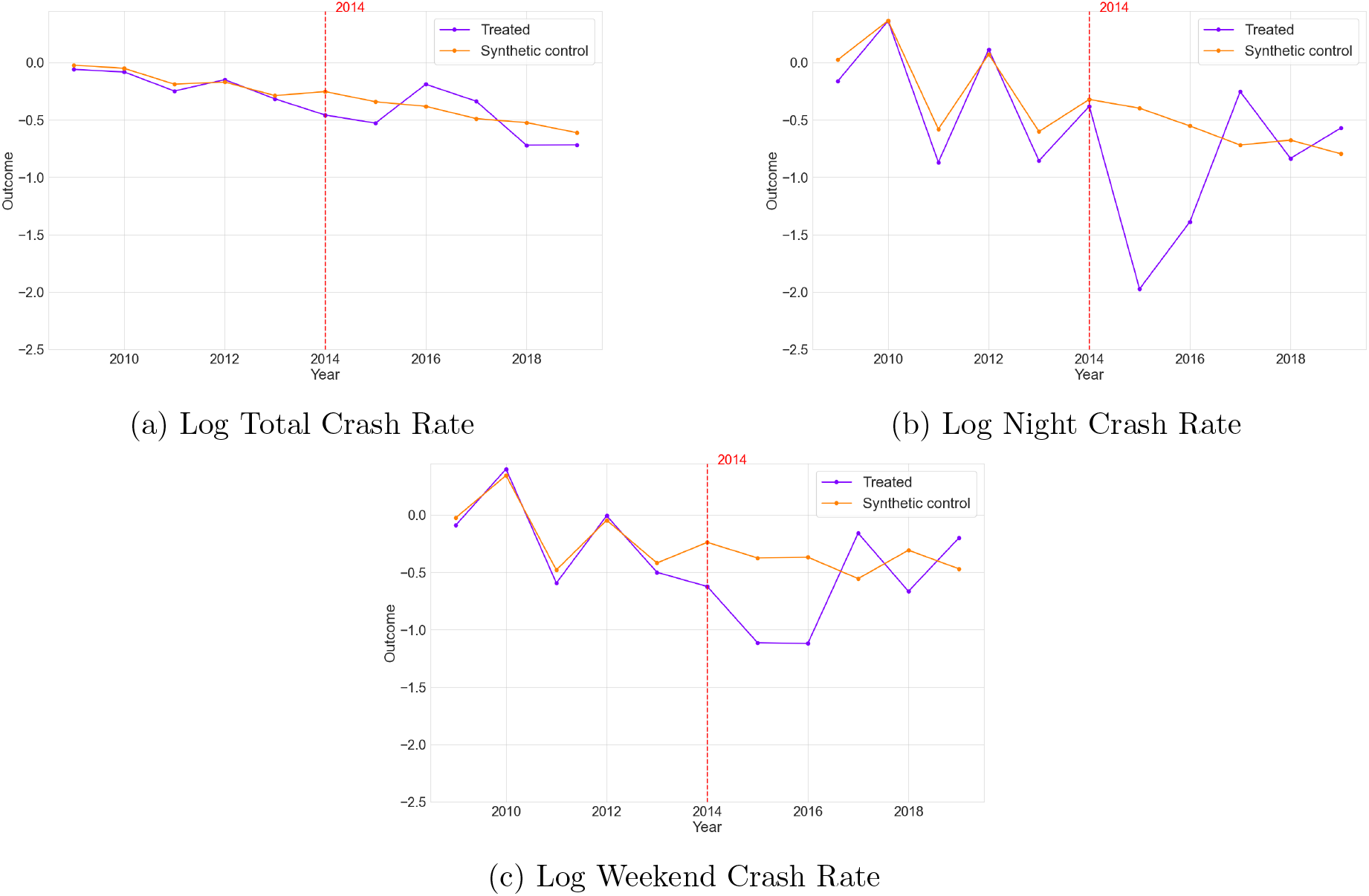
Observed and synthetic crash trends before and after BAC policy implementation: Heterogeneous effects, Cluster 2

**Figure 7:**
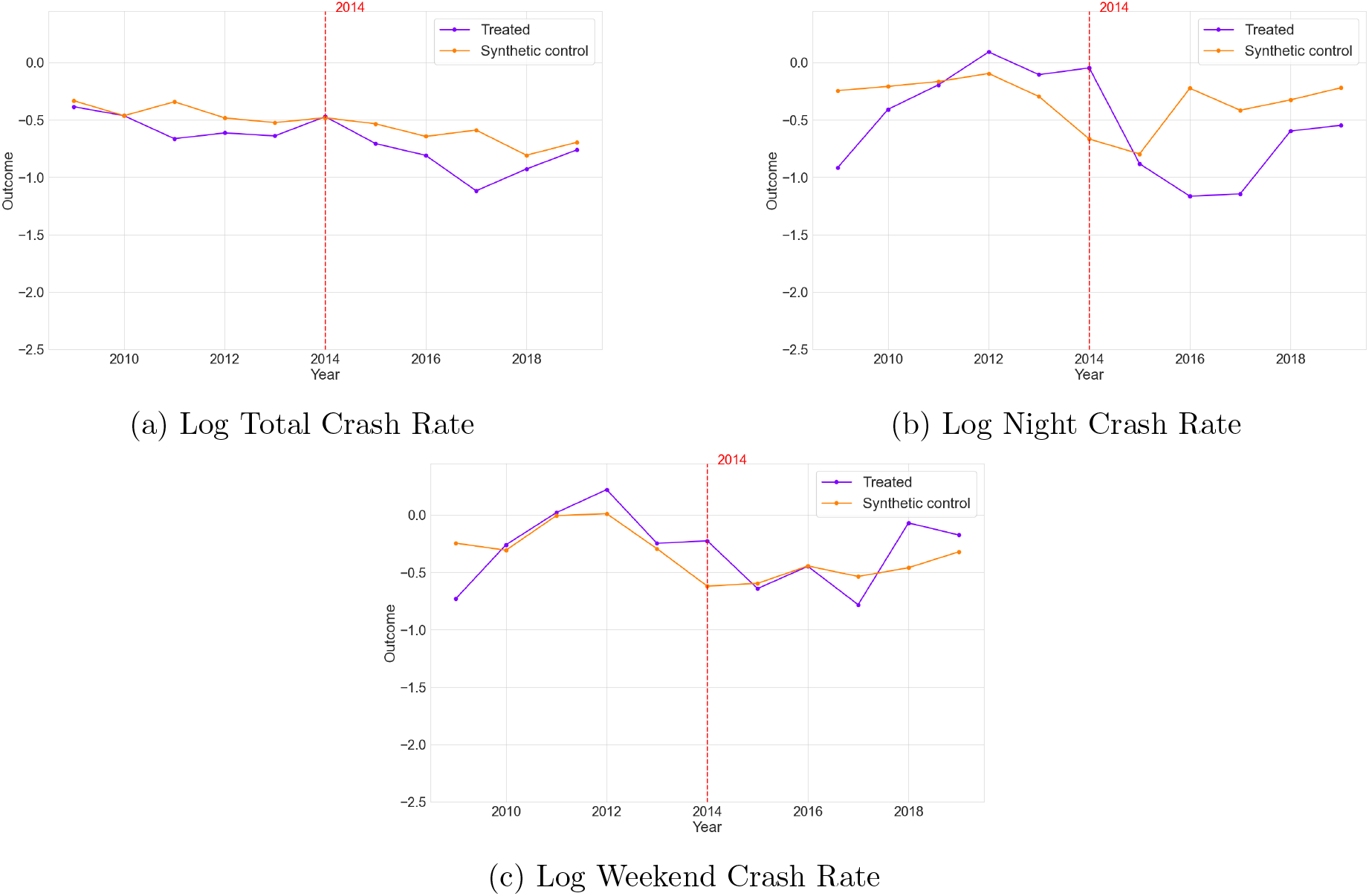
Observed and synthetic crash trends before and after BAC policy implementation: Heterogeneous effects, Cluster 3

**Figure 8:**
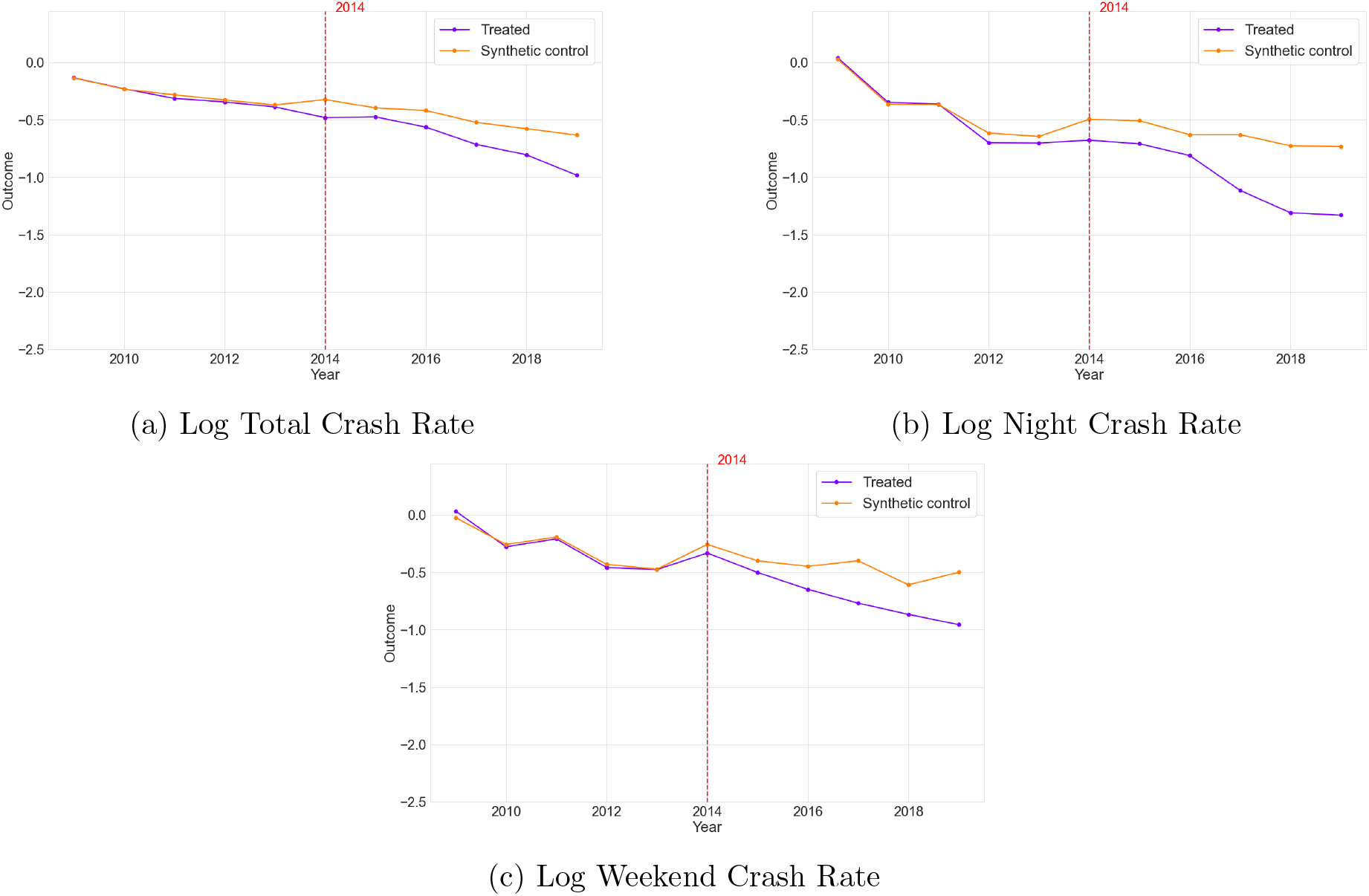
Observed and synthetic crash trends before and after BAC policy implementation: Heterogeneous effects, Cluster 4

## 4 Discussion

### 4.1 Principal Findings

This study evaluates the impact of Scotland’s 2014 reduction in the legal blood alcohol concentration (BAC) limit using a Synthetic Difference-in-Differences (SDID) design applied to a national panel of small-area data from Great Britain over the period 2008 to 2019. Overall, the results indicate that the reform led to reductions in road traffic crash rates.

At the aggregate level, the estimated effects correspond to reductions of about 12% in total crash rates, 15.6% in night-time crash rates, and 12.4% in weekend night-time crash rates. The larger reductions observed for night-time and weekend night-time crashes, which are more closely linked to alcohol-related driving risk, are consistent with the interpretation that the reform primarily affected drink-driving behaviour rather than road traffic risk more generally.

The analysis also indicates substantial heterogeneity in policy effects across area types. The largest reductions are observed in large rural road-network areas and typical rural areas, where the estimated declines in total crash rates exceed 16%, and night-time crash reductions reach almost 48% in some cases. Moderate but statistically significant reductions are also observed in dense urban areas. By contrast, estimated effects are smaller and not statistically distinguishable from zero in suburban/semi-urban areas and urban–rural transition areas. Taken together, these findings suggest that the effects of stricter BAC limits are context-dependent and that analyses based only on aggregate average effects may conceal important variation in how such reforms operate across space.

The stronger reductions observed in rural and less densely populated areas are intuitively plausible. In these settings, alcohol-related travel may be more car-dependent because public transport, taxis, and walkable alternatives are often less available, particularly at night. As a result, a reduction in the legal BAC limit may have a more direct influence on the decision to drive after drinking. Rural crashes may also involve longer trip distances and higher-speed roads, meaning that behavioural changes among drink-drivers could translate into larger reductions in observed crash rates. By contrast, the weaker and statistically uncertain effects in suburban, semi-urban, and urban–rural transition areas may reflect more mixed travel patterns, lower baseline alcohol-related crash risk, or less direct exposure to the specific behaviour targeted by the reform. These explanations should be interpreted as hypotheses rather than definitive mechanisms, but they help explain why the estimated effects are spatially uneven rather than uniform across Scotland.

### 4.2 Strengths and Limitations of the Study

This study has several strengths. First, it uses a large national panel covering all police-reported road traffic collisions in Great Britain over more than a decade. Second, the analysis is conducted at a fine spatial scale, allowing policy impacts to be examined at the level of small areas rather than only at the level of countries or local authorities. Third, the SDID framework provides a more credible counterfactual than a standard unweighted difference-in-differences design by improving pre-treatment comparability between treated and control areas through weighting. Fourth, the study considers both aggregate and heterogeneous effects, thereby providing a more nuanced understanding of how the BAC reform operated across different types of areas.

Several limitations should also be considered. First, the analysis relies on police-reported collision data, which may be affected by under-reporting or changes in reporting practices over time. Although there is no clear reason to expect such changes to differentially affect Scotland relative to England and Wales, some measurement error may remain. Second, the analysis uses population as a proxy for exposure because comparable traffic-volume data are not available consistently at the same spatial scale. Population-standardised crash rates, therefore, may not fully capture differences in underlying road use, particularly in areas with substantial commuting, tourism, or through traffic.

Third, the analysis uses annual data, whereas the reform came into force in December 2014. In the main specification, 2014 is treated as the intervention year because behavioural responses, public awareness, and enforcement-related changes may begin around the announcement and rollout of the reform rather than only after formal implementation. Nonetheless, annual aggregation may still blur the precise timing of behavioural adjustment. Sensitivity analyses that instead treat 2015 as the start of the post-intervention period yield qualitatively similar results, suggesting that the main findings are not driven by the treatment-timing convention.

Fourth, although SDID improves causal identification, its validity still depends on the assumption that no other major Scotland-specific changes affecting road safety occurred contemporaneously with the BAC reform in a way that would materially bias the estimates. Spillover effects, including cross-border behavioural or enforcement responses, are also difficult to rule out completely. Finally, the heterogeneity analysis is based on clustering area characteristics. While this provides a useful way to examine conditional treatment effects across similar types of places, cluster-specific estimates are subject to lower statistical power and should therefore be interpreted with appropriate caution.

### 4.3 Comparison with Other Studies

The present findings differ from earlier evaluations of Scotland’s 2014 BAC reform, which generally reported limited or no statistically significant effects on road traffic collisions. For example, Haghpanahan et al. [29], using a comparative interrupted time-series design with weekly country-level data from 2013 to 2016, found no detectable reduction in crash outcomes following the policy change. Similarly, Francesconi and James [28], using monthly local-authority-level data from 2009 to 2016, reported no statistically significant impact and suggested that weak enforcement and limited behavioural responses may have constrained the policy’s effectiveness.

Several differences in study design may help explain why the present analysis reaches different conclusions. First, this study uses a longer post-treatment period, extending to 2019, which allows greater scope for delayed behavioural responses to emerge. Second, the analysis is conducted at a finer spatial resolution, making it possible to detect heterogeneity across areas that may be obscured in more aggregated designs. Third, the SDID framework provides a different approach to identification from conventional difference-in-differences, improving pre-treatment comparability through weighting and potentially yielding a more credible counterfactual when treated and control areas differ in underlying trends. Fourth, the use of time-based proxy outcomes for alcohol-related risk may capture relevant behavioural changes more consistently than outcomes derived from recorded breath-test results, which may be affected by incomplete testing and reporting.

More broadly, the results are in line with the wider international literature suggesting that lower legal BAC limits can reduce alcohol-related collisions and injuries. In particular, the larger estimated effects during night-time and weekend night-time periods are consistent with evidence that such policies are most likely to affect collisions occurring during periods of elevated drink-driving risk. The present study also extends the literature by showing that these effects may vary markedly across spatial contexts, with stronger reductions in rural and less densely populated areas than in suburban or transitional settings.

### 4.4 Meaning of the Study for Clinicians and Policy Makers

These findings suggest that lowering the legal BAC limit can contribute meaningfully to road safety improvement, particularly during periods and in locations where alcohol-related driving risk is greatest. In the context of ongoing policy debate in England, Wales, and Northern Ireland, the results provide evidence that Scotland’s reform was associated with reductions in crash rates rather than with null effects across the board.

At the same time, the results indicate that the impact of BAC policy is not spatially uniform. The stronger estimated effects in rural and less densely populated areas are consistent with the idea that the reform had the greatest marginal impact where alcohol-related driving risk was most directly linked to car dependency. In areas with fewer late-night transport alternatives, the lower BAC limit may have changed the perceived acceptability or risk of driving after drinking more strongly than in places where alternative travel options were more available. In dense urban areas, the reform may still have reduced crashes, but the marginal effect may be smaller because individuals already had more non-driving options after alcohol consumption. The weaker effects in suburban and transition areas may reflect more heterogeneous travel behaviour and lower statistical power within these clusters. These patterns suggest that national BAC reforms may be most effective when supported by locally tailored enforcement, communication, and transport measures.

### 4.5 Unanswered Questions and Future Research

This study focuses on the overall effect of Scotland’s BAC reform on crash outcomes, but it is less able to isolate the specific pathways through which the reform operated. In particular, changes in enforcement intensity, publicity, perceptions of apprehension risk, and the availability of alternative transport may all mediate the relationship between stricter legal limits and crash outcomes. Because some of these factors may themselves respond to the reform, they cannot be straightforwardly included as controls without risking post-treatment bias. Future research should therefore examine whether the observed effects are mediated by specific enforcement and behavioural mechanisms, and whether these mechanisms differ systematically across area types.

Further work is also needed to explore outcome measures that more directly reflect alcohol-related crash involvement. The present study relies on time-based proxy outcomes because directly recorded alcohol-related crashes are not publicly available in STATS19 and may, in any case, be affected by incomplete testing and reporting. Although the larger estimated effects for night-time and weekend night-time crashes are consistent with an alcohol-related mechanism, future studies using linked health, police, or toxicology data could help clarify this relationship more directly.

Finally, the study period ends in 2019 because the COVID-19 pandemic introduced major disruptions to traffic volumes, travel behaviour, enforcement patterns, and broader social activity, creating a structural break that complicates causal identification. Future research should revisit the longer-term effects of Scotland’s BAC reform once a sufficiently long and stable post-pandemic period becomes available.

## 5 Conclusions

This study evaluates the impact of Scotland’s 2014 reduction in the legal blood alcohol concentration (BAC) limit using a Synthetic Difference-in-Differences (SDID) framework applied to small-area data across Great Britain from 2008 to 2019. The findings indicate that the reform led to reductions in road traffic crash rates, with estimated declines of about 12% in total crashes, 15.6% in night-time crashes, and 12.4% in weekend night-time crashes.

The results also show substantial spatial heterogeneity in policy effects. Larger reductions are observed in rural and less densely populated areas, particularly during higher-risk periods linked to alcohol-related driving, whereas effects are smaller or not statistically distinguishable from zero in suburban and transitional areas. These patterns suggest that the effects of BAC policy are context dependent and may vary with local transport conditions, enforcement environments, and behavioural responses.

By combining a robust causal inference framework with high-resolution spatial data, this study provides new evidence on the effects of Scotland’s BAC reform and complements previous evaluations based on more aggregated data and shorter follow-up periods. Overall, the findings support the view that lower legal BAC limits can contribute to improved road safety, while also highlighting the importance of accounting for local context when designing and evaluating such interventions.

## Data Availability

All data produced are open-access and available online at the links provided in the manuscript.

## APPENDIX A Technical Supplement to the Methods Section

This appendix provides additional technical details on the implementation of the Synthetic Difference-in-Differences (SDID) estimator and the clustering procedure used in the heterogeneous treatment effect analysis.

### A.1 Why SDID is preferred

Among the available extensions to Difference-in-Differences (DiD) and synthetic control methods, Synthetic Difference-in-Differences (SDID) is particularly well suited to the present setting. Compared with conventional DiD, SDID reduces reliance on an unweighted parallel-trends comparison by constructing a more comparable control counterfactual through unit weighting, while time weighting further emphasises pre-treatment periods that are most informative for the post-treatment counterfactual. Compared with conventional synthetic control methods (SCM), SDID is naturally adapted to panel settings with many treated units observed over time, rather than a single treated unit and a donor pool.

Related approaches, including demeaned or intercept-shift SCM [64], augmented SCM [65], generalised SCM [66], and matrix completion methods [67], offer useful alternatives for addressing imperfect pre-treatment fit or latent factor structure. However, these approaches rely more heavily on model-based extrapolation or are less directly targeted to a simultaneous, multiple-treated-unit design. SDID therefore provides an appropriate balance between flexibility and transparency: it preserves the weighting-based logic of synthetic control while embedding it within a DiD framework. This is especially attractive here, where treated and control areas are numerous, pre-policy trajectories are heterogeneous, and the credibility of the counterfactual comparison is central to identification.

### A.2 Estimation of unit and time weights

The SDID estimator determines unit and time weights through separate regularised optimisation problems designed to align pre-treatment trends. Unit weights are obtained by solving:

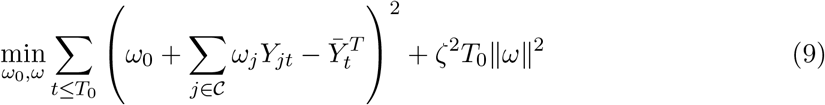

subject to

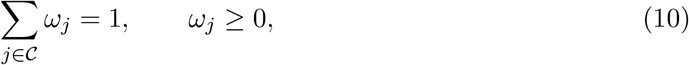

where 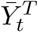 denotes the average outcome for treated units at time *t*, and *ζ* is a regularisation parameter that limits overfitting.

Time weights are estimated as:

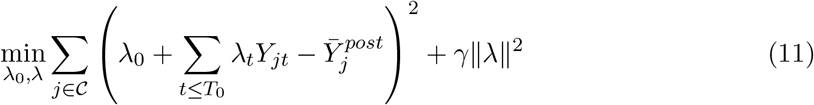

subject to

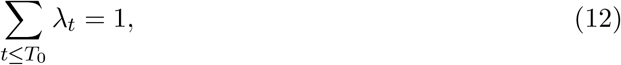

where 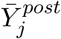 denotes the average post-treatment outcome for control unit *j*, and *γ* is a regularisation parameter. These optimisation problems ensure that the synthetic control group reproduces the pre-intervention trajectory of the treated units while also adjusting for time-varying shocks common across units.

### A.3 Placebo-based inference

In each placebo iteration, a subset of control units equal in size to the treated group is randomly selected from the donor pool without replacement and assigned placebo treatment status. The SDID model is then re-estimated using this placebo assignment, yielding a placebo treatment effect 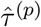. Repeating this procedure *P* = 300 times generates an empirical distribution of placebo effects under the null hypothesis of no treatment effect. Because the BAC-limit reduction was expected to reduce crash rates, statistical significance was assessed using a one-sided lower-tail placebo p-value, defined as the proportion of placebo estimates less than or equal to the observed SDID estimate, with a finite-sample correction. The placebo-based p-value is computed as:

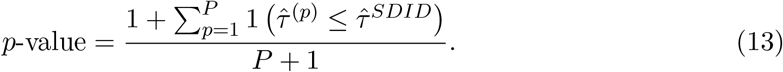

The sampling variability of the estimator is approximated using the empirical distribution of placebo treatment effects. Specifically, the variance of the estimator is computed as:

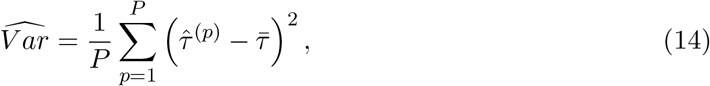

where

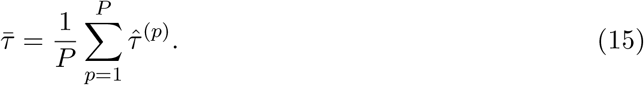

The corresponding standard error is defined as:

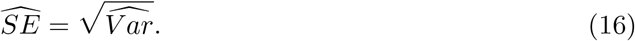

While placebo-based p-values provide a non-parametric measure of statistical significance, confidence intervals are constructed using a normal approximation based on the empirical variance of placebo estimates. This provides a practical summary of uncertainty from the placebo distribution and is commonly used in SDID applications. Ninety-five per cent confidence intervals are constructed as:

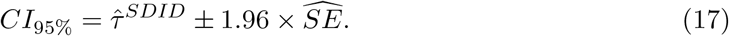

This placebo-based approach is well-suited to panel settings with serial correlation and complex dependence structures, where conventional variance estimators may be unreliable.

### A.4 Principal component analysis

Principal Component Analysis (PCA) is used as a dimensionality-reduction step before clustering because the pre-treatment area characteristics are high-dimensional and correlated. PCA is applied to the standardised clustering variables, and the first seven principal components are retained. These components jointly explain at least 90% of the total variance in the clustering variables (Figure 9).

**Figure 9:**
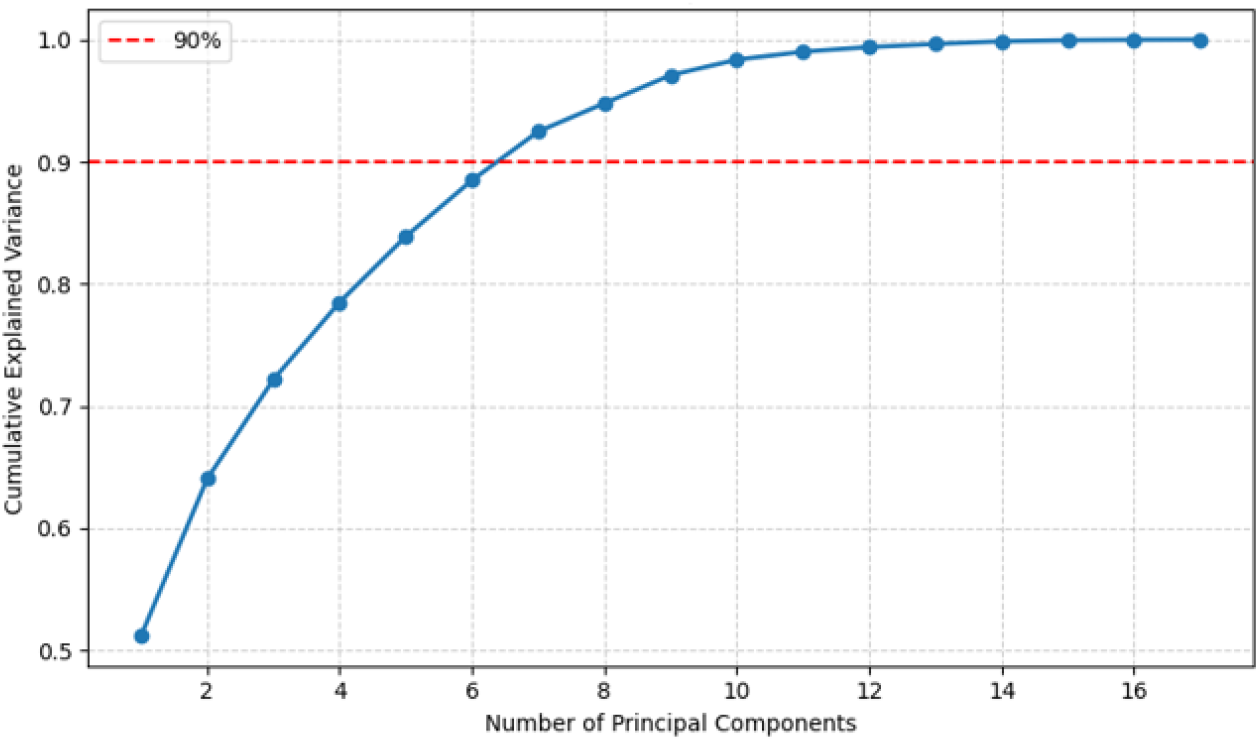
Cumulative explained variance from principal component analysis

### A.5 Clustering implementation and validation

Clustering is performed on the retained PCA scores using hierarchical agglomerative clustering with Ward’s linkage. Let *d*(*A, B*) denote the distance between clusters *A* and *B*. Ward’s method merges the pair of clusters that minimises the increase in within-cluster sum of squares at each step, thereby favouring compact and internally homogeneous clusters. This property is particularly useful in the present setting, where area types may differ in both size and composition.

The number of clusters is selected using two standard validation criteria. The silhouette score for observation *i* is defined as:

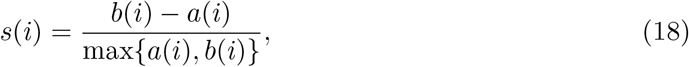

where *a*(*i*) is the average distance between observation *i* and all other observations in its own cluster, and *b*(*i*) is the minimum average distance between observation *i* and observations in another cluster. Higher average silhouette scores indicate better-defined cluster separation.

The Davies–Bouldin index is defined as:

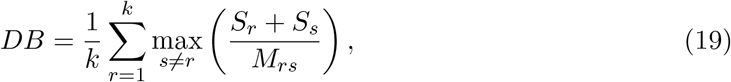

where *S*_*r*_ and *S*_*s*_ denote within-cluster dispersion for clusters *r* and *s*, and *M*_*rs*_ denotes the distance between cluster centroids. Lower values indicate better separation relative to within-cluster dispersion.

Based on these criteria, and subject to the requirement that each cluster contains a sufficient number of treated and control units for estimation, the number of clusters is set to *k* = 5.

### A.6 Cluster profiles

Figure 10 reports cluster profiles based on standardised (z-scored) values of the variables used in the clustering procedure.

**Figure 10:**
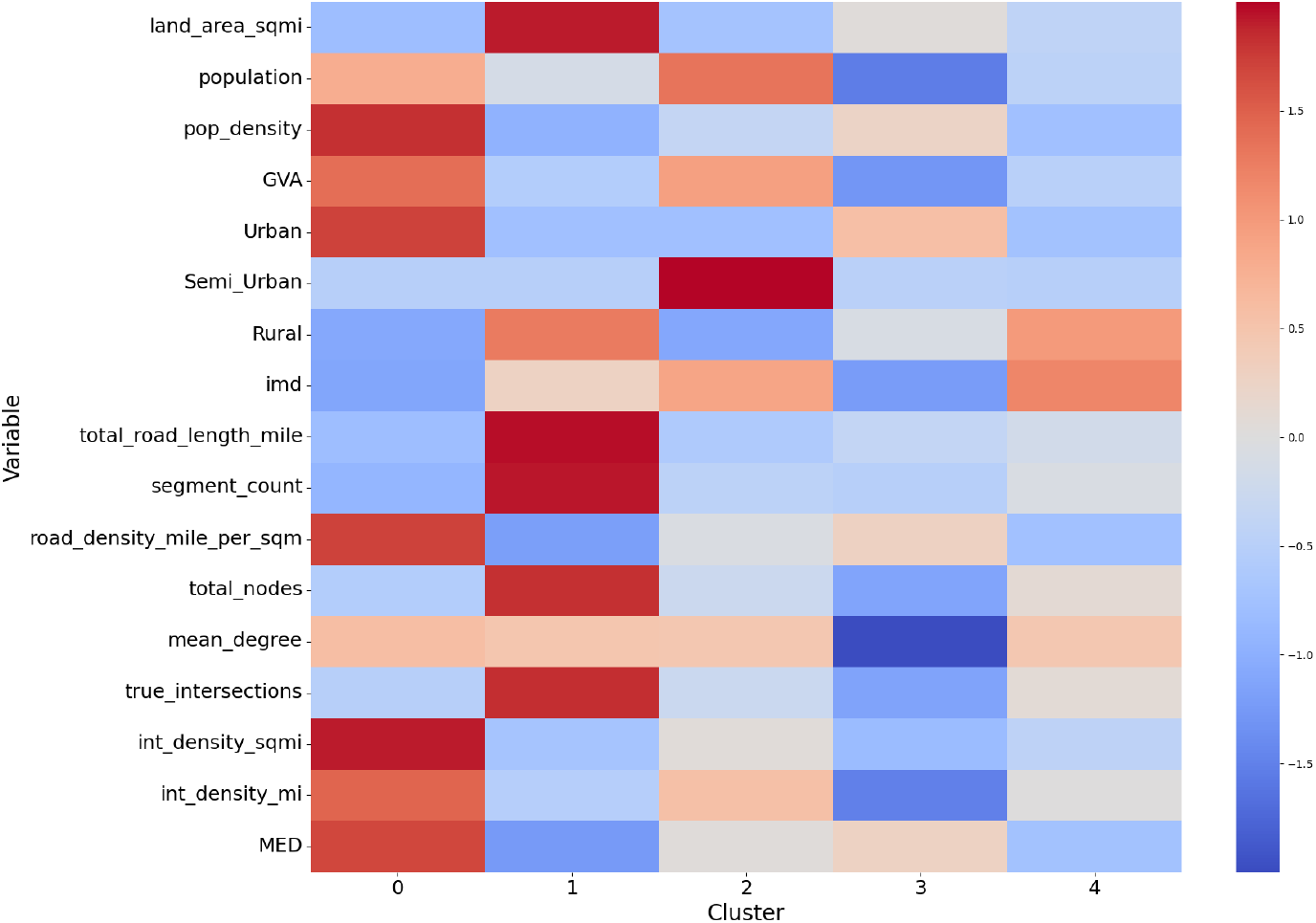
Hierarchical cluster profiles (z-scored)

A cluster profile represents the average value of each variable within a cluster. Positive values indicate above-average levels and negative values indicate below-average levels relative to the full sample. These profiles highlight clear differences across clusters in terms of population size, density, economic activity (GVA), road-network structure, and accessibility (MED), and provide the basis for the cluster labels reported in the main text.

### A.7 Pre-treatment diagnostics within clusters

To assess whether treated and control units are sufficiently comparable within each cluster before the policy intervention, we examine pre-treatment outcome trajectories separately for each cluster and each outcome measure. Figure 11 presents these pre-treatment trends for treated and control units within each cluster across the three outcomes.

**Figure 11:**
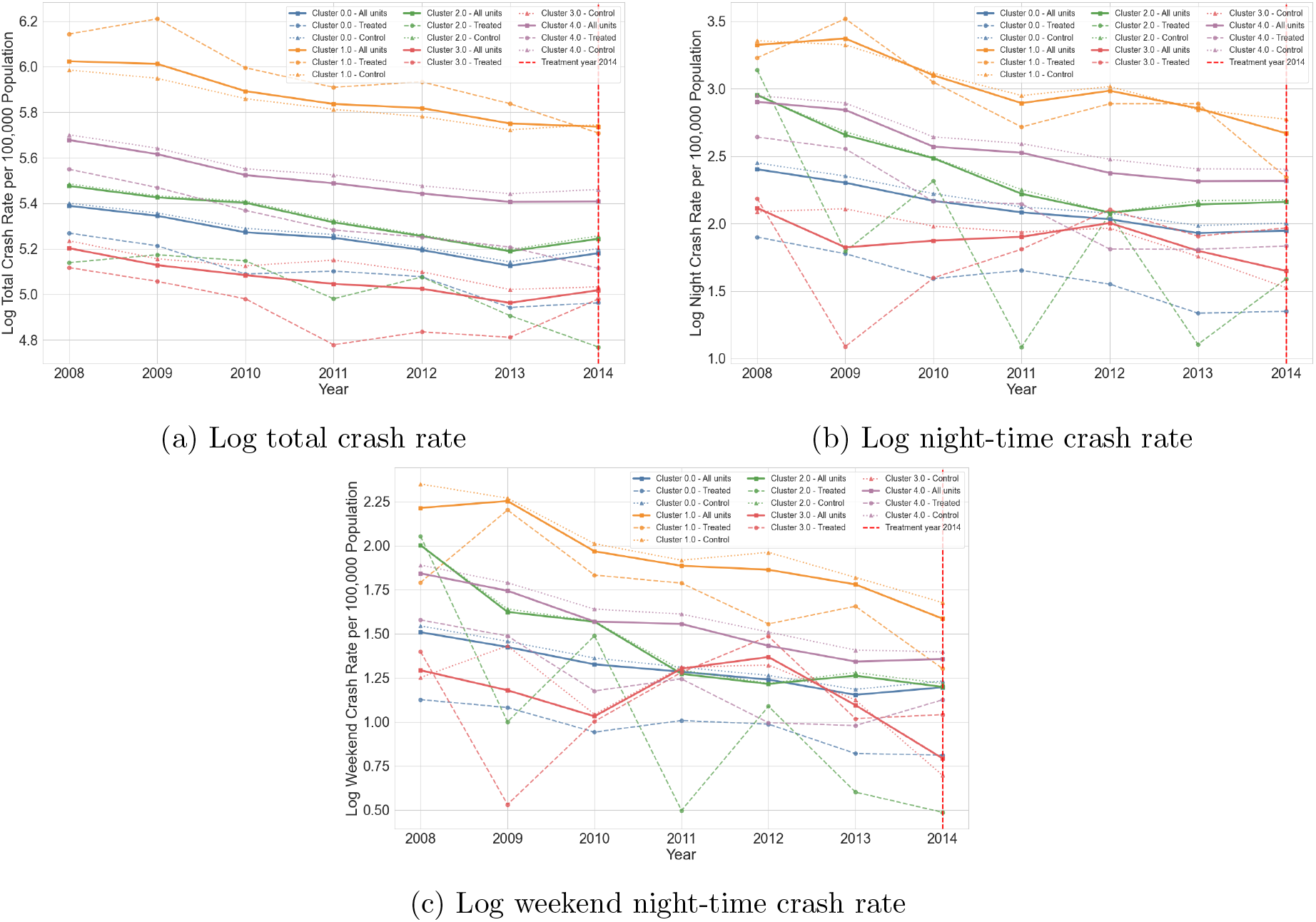
Pre-treatment trends in treated and control units within each cluster

**Figure 12:**
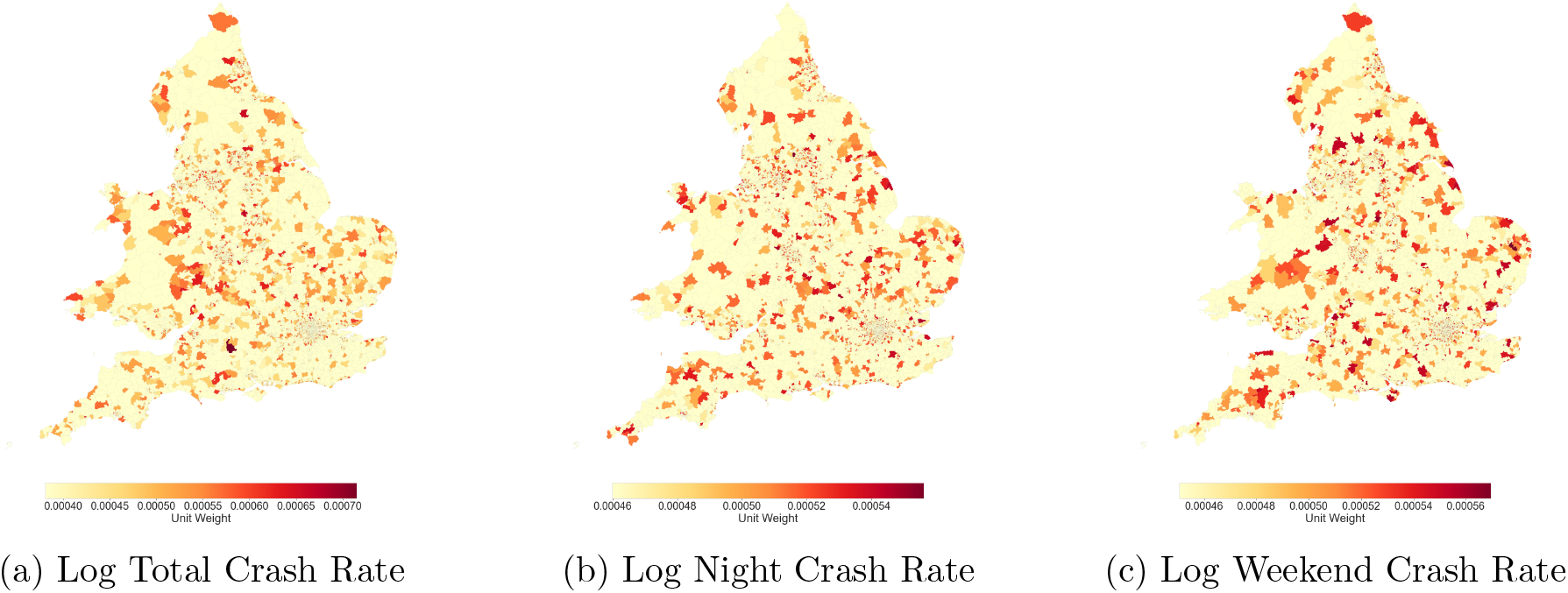
Unit weights across outcomes in the full sample

**Figure 13:**
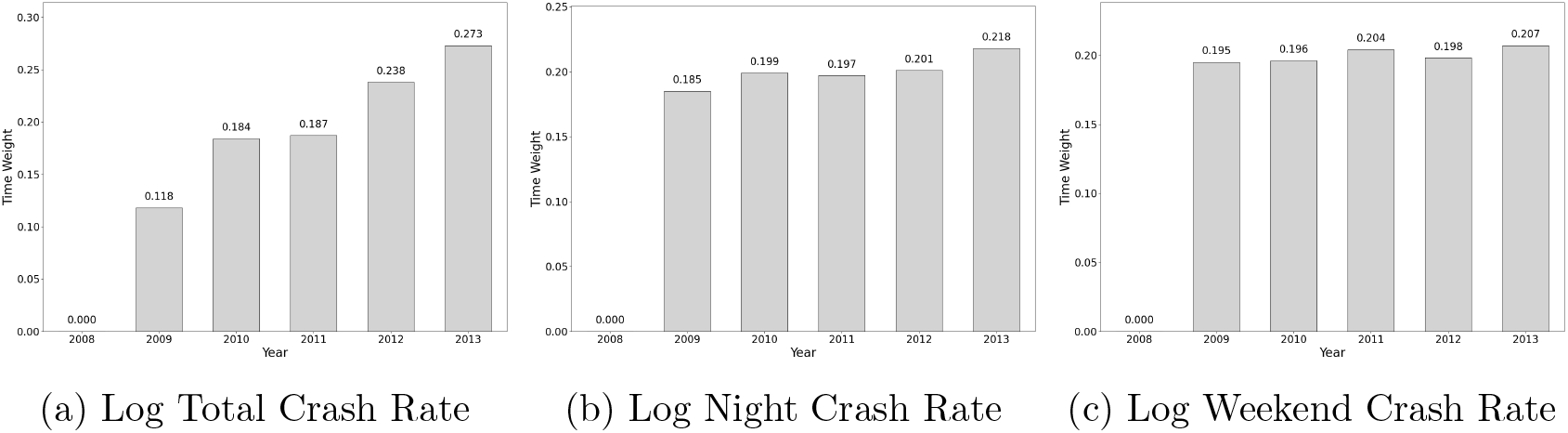
Time weights across outcomes in the full sample

**Figure 14:**
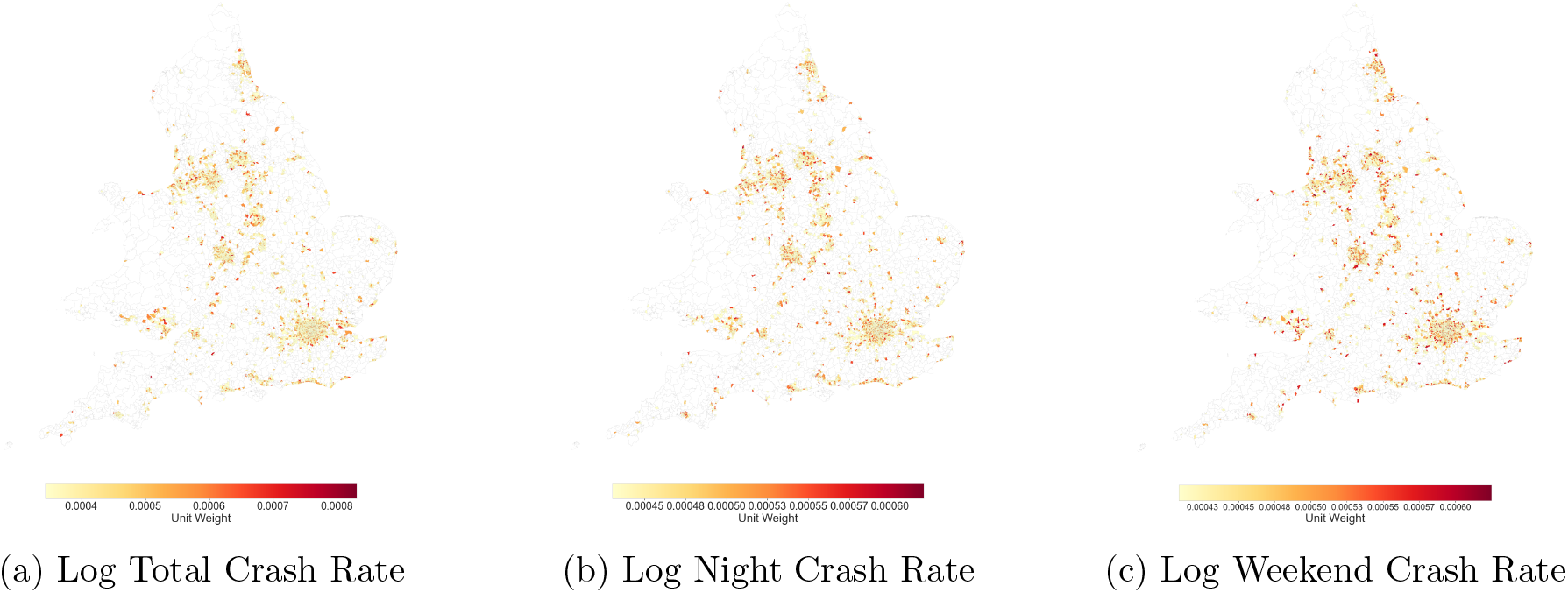
Unit weights across outcomes in Cluster 0

**Figure 15:**
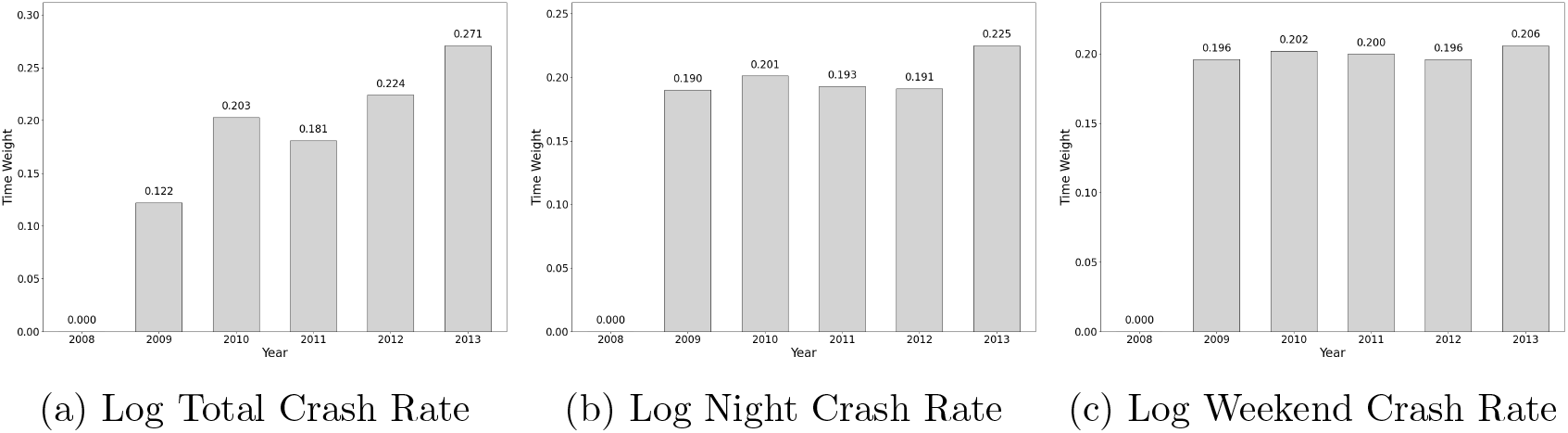
Time weights across outcomes in Cluster 0

**Figure 16:**
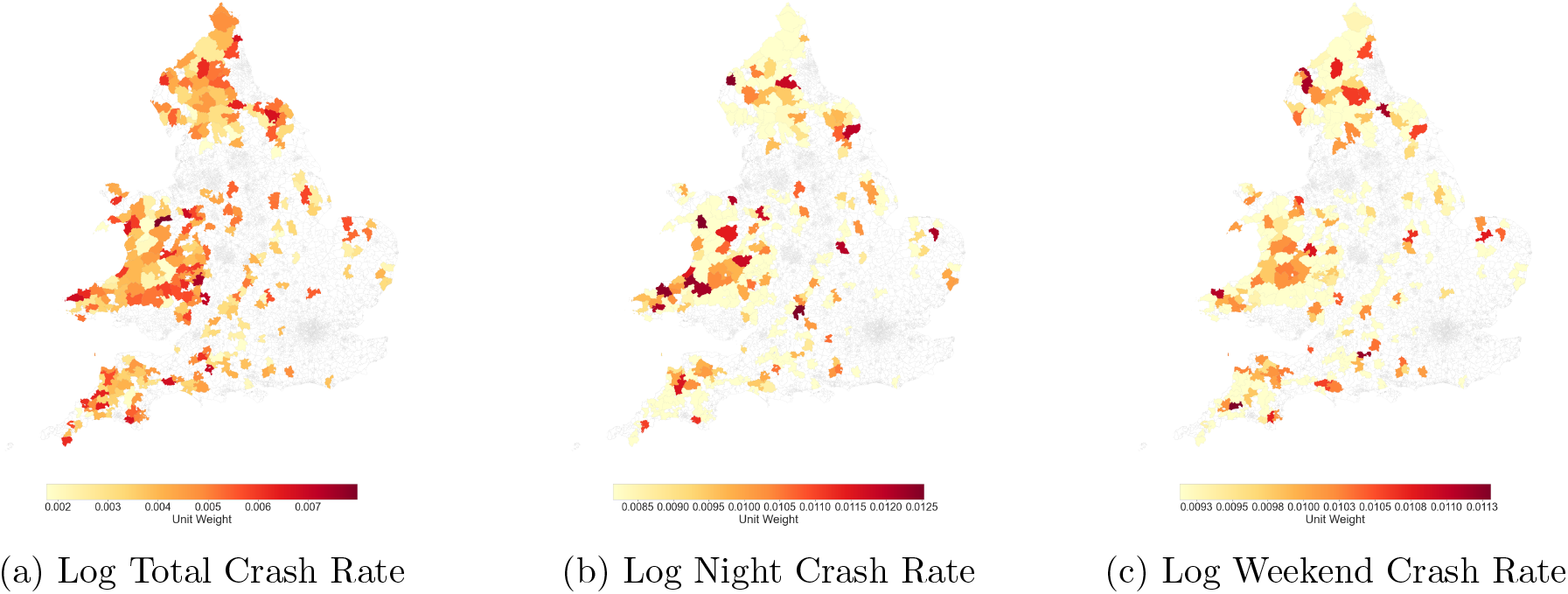
Unit weights across outcomes in Cluster 1

**Figure 17:**
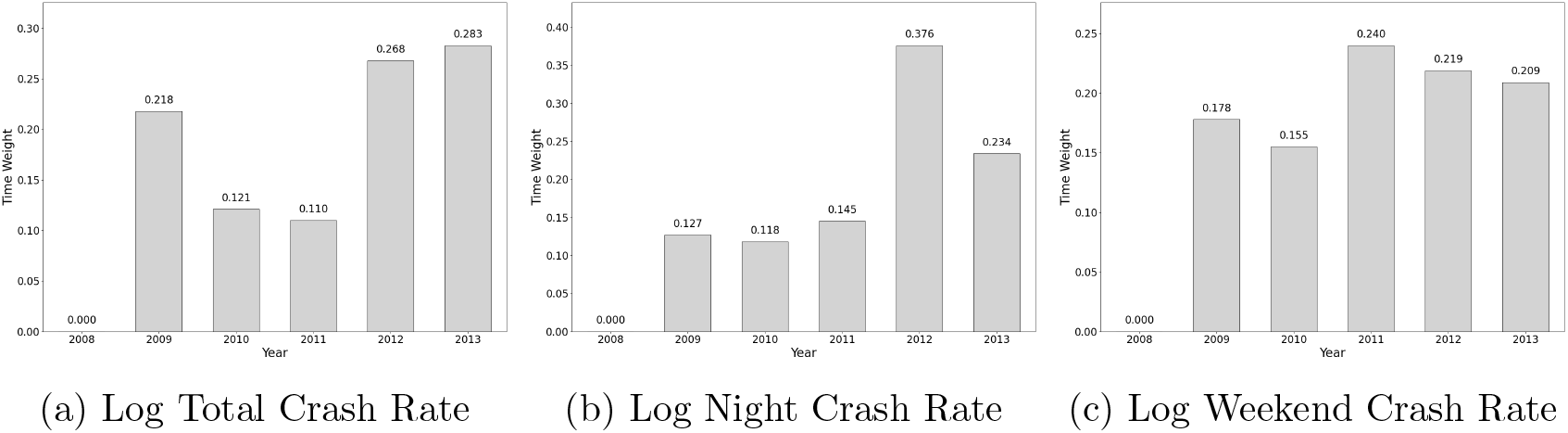
Time weights across outcomes in Cluster 1

**Figure 18:**
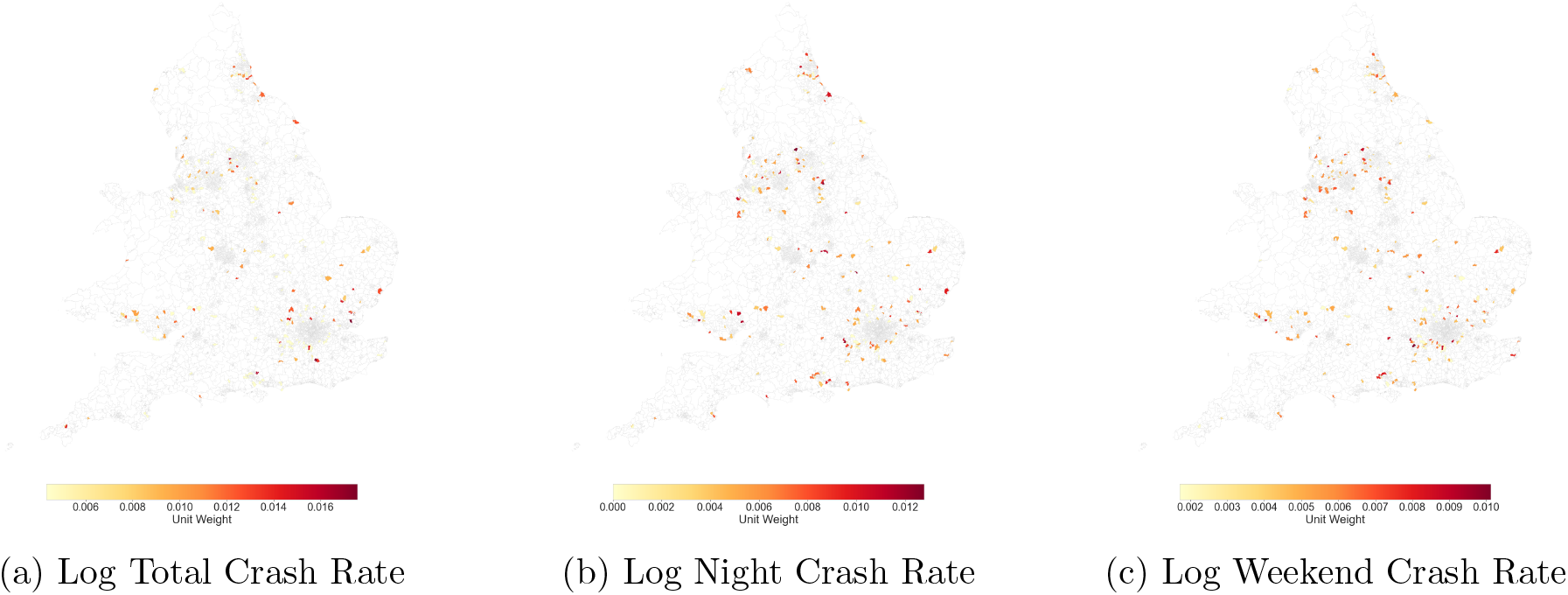
Unit weights across outcomes in Cluster 2

**Figure 19:**
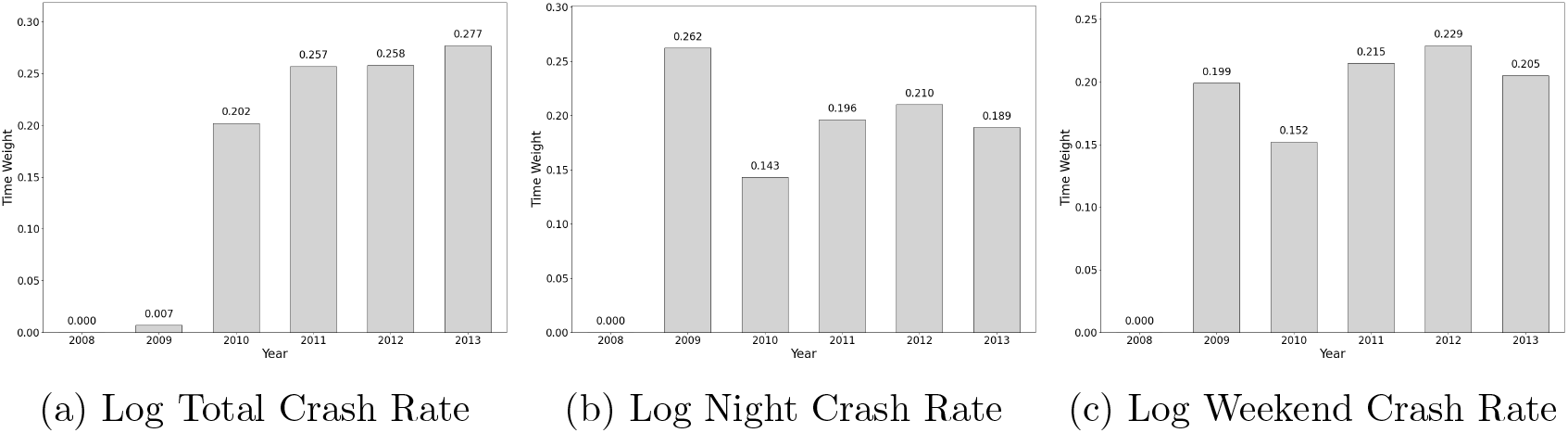
Time weights across outcomes in Cluster 2

**Figure 20:**
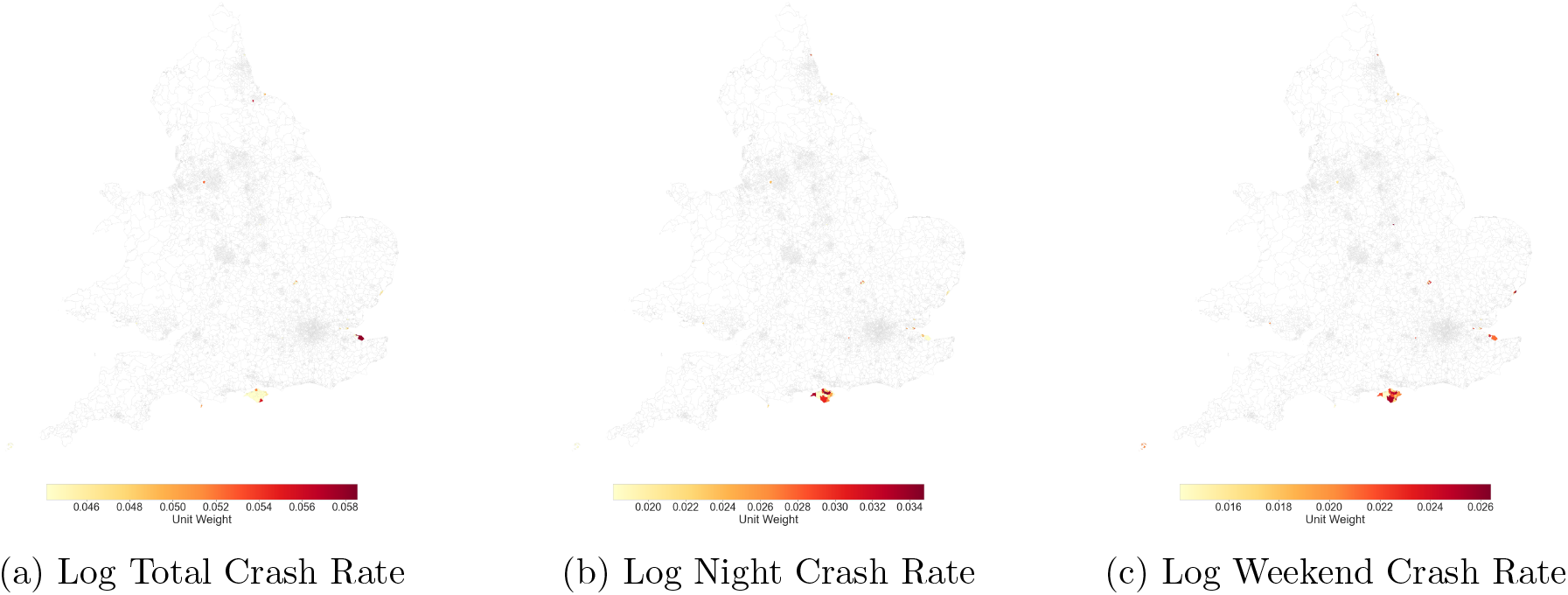
Unit weights across outcomes in Cluster 3

**Figure 21:**
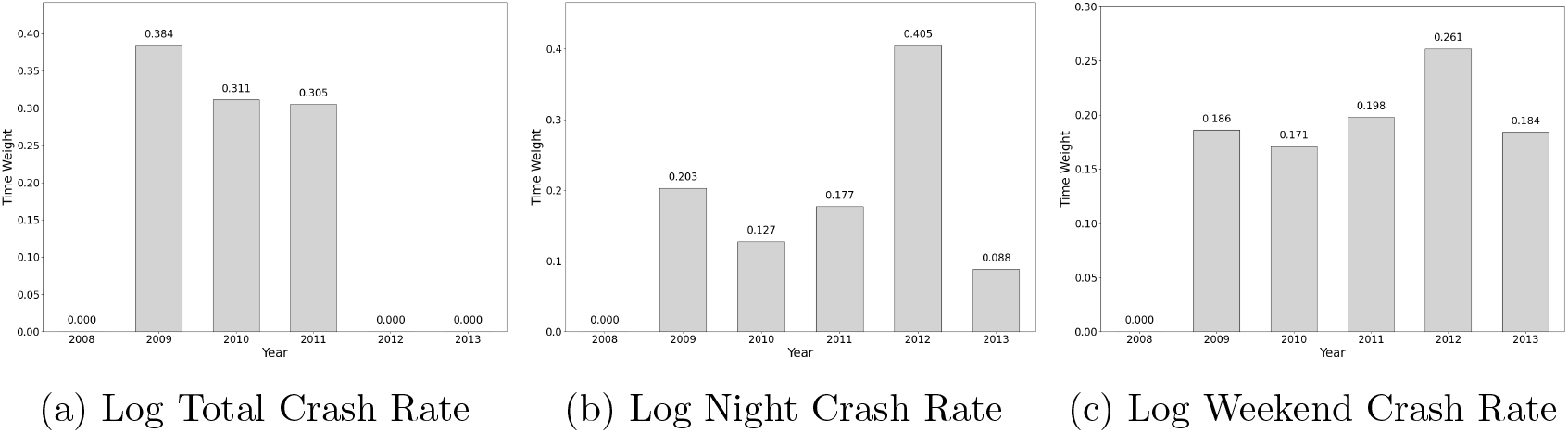
Time weights across outcomes in Cluster 3

**Figure 22:**
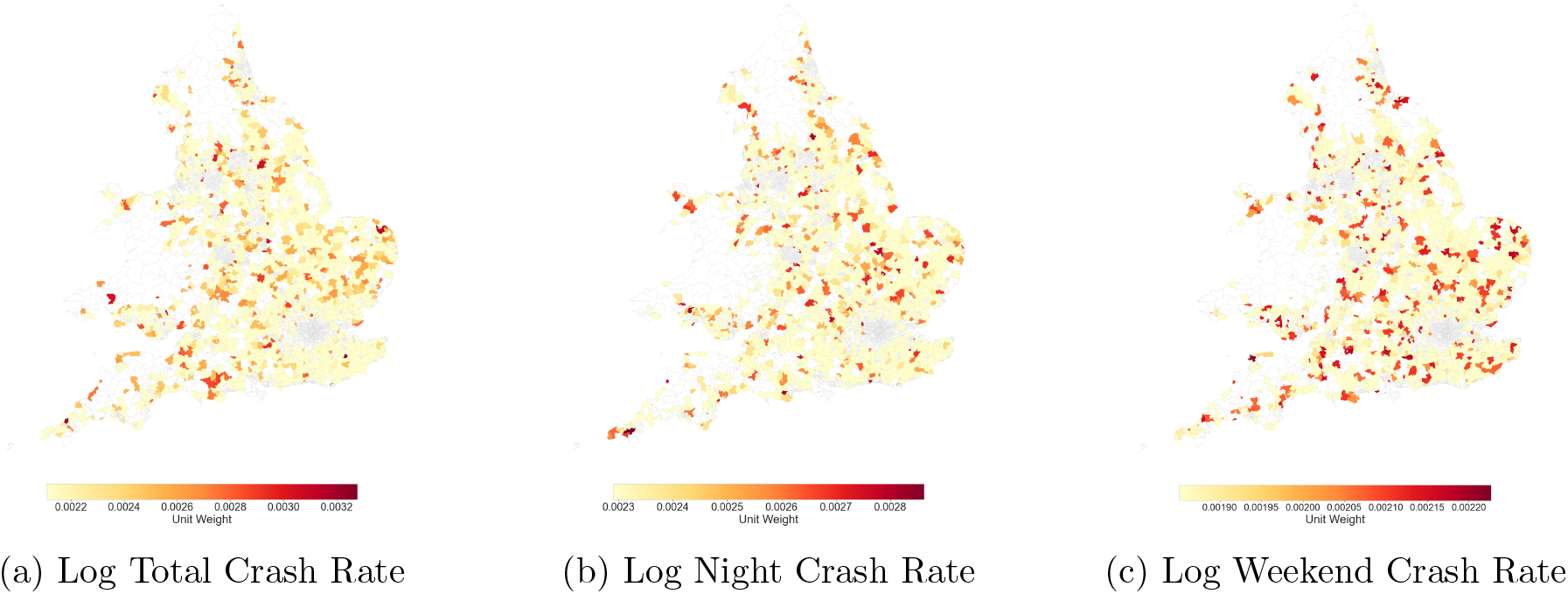
Unit weights across outcomes in Cluster 4

**Figure 23:**
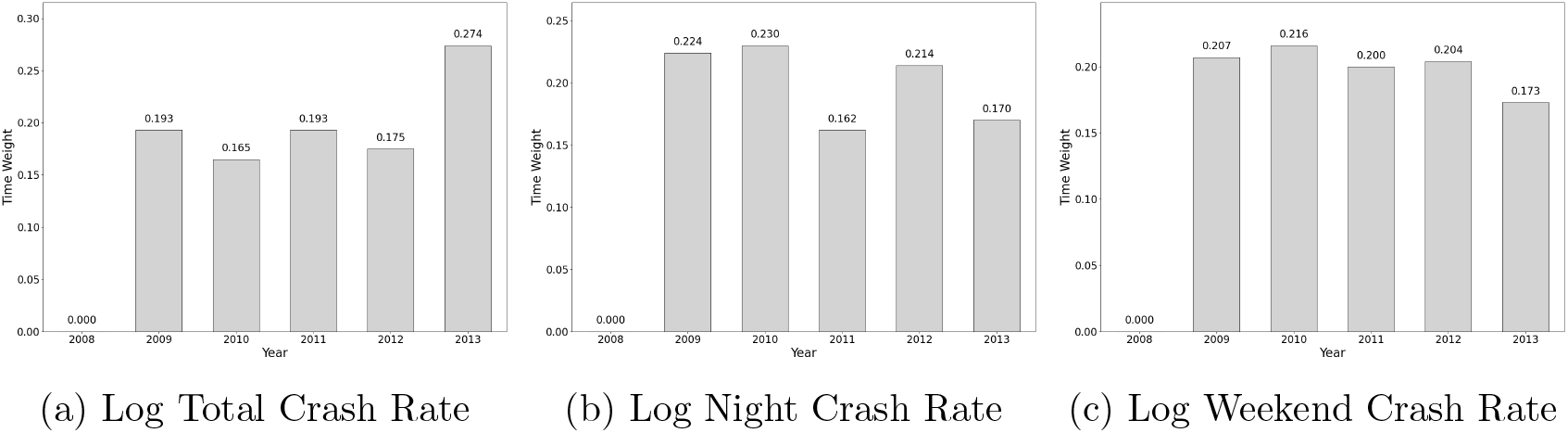
Time weights across outcomes in Cluster 4

Overall, the trajectories indicate a reasonable degree of comparability between treated and control areas prior to the implementation of the BAC policy. For the total crash rate outcome, treated and control units generally exhibit similar declining trends across most clusters, although some level differences remain in certain groups. Night-time and weekend night-time crash outcomes display greater year-to-year variability, particularly in clusters with smaller numbers of treated units, reflecting the lower frequency of these events. Nevertheless, the overall direction and temporal evolution of the trends remain broadly comparable between treated and control units within clusters, providing supporting evidence for the use of SDID within clusters.

## APPENDIX B Sensitivity Analysis: Treatment Year as 2015

## APPENDIX C Unit and Time Weights

### C.1 Full Sample

### C.2 Cluster 0

### C.3 Cluster 1

### C.4 Cluster 2

### C.5 Cluster 3

### C.6 Cluster 4

## APPENDIX D Placebo-Based Inference Histograms

Figures 24-29 compare the placebo tau distribution with the estimated treatment-effect distribution. The placebo distribution was obtained from the permutation procedure, while the real tau distribution was approximated using the estimated treatment effect and its standard error. The vertical dotted lines show the 95% confidence interval, and the dashed black line represents the zero-effect reference.

**Figure 24:**
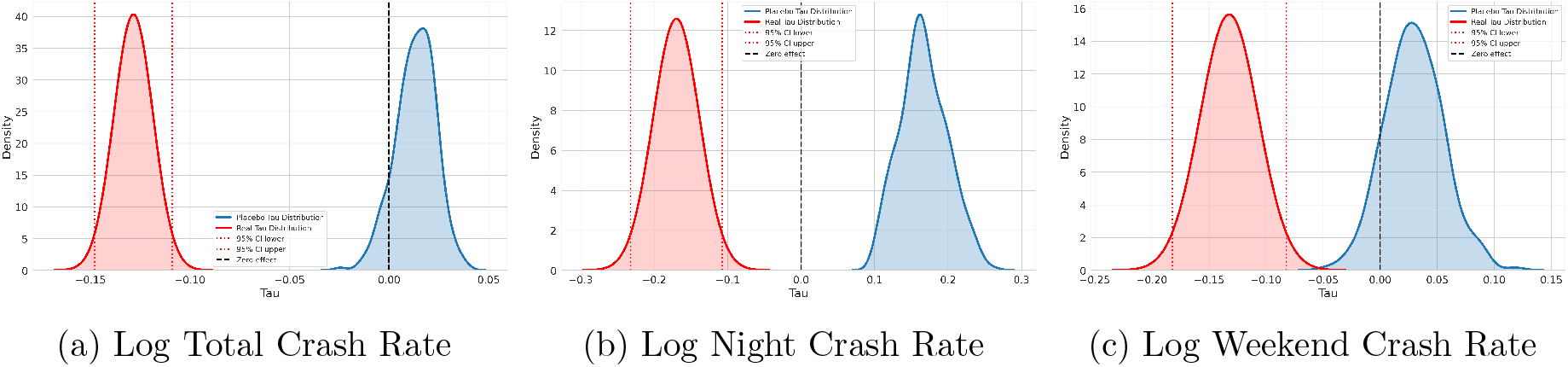
Real and placebo tau distributions in the full sample

**Figure 25:**
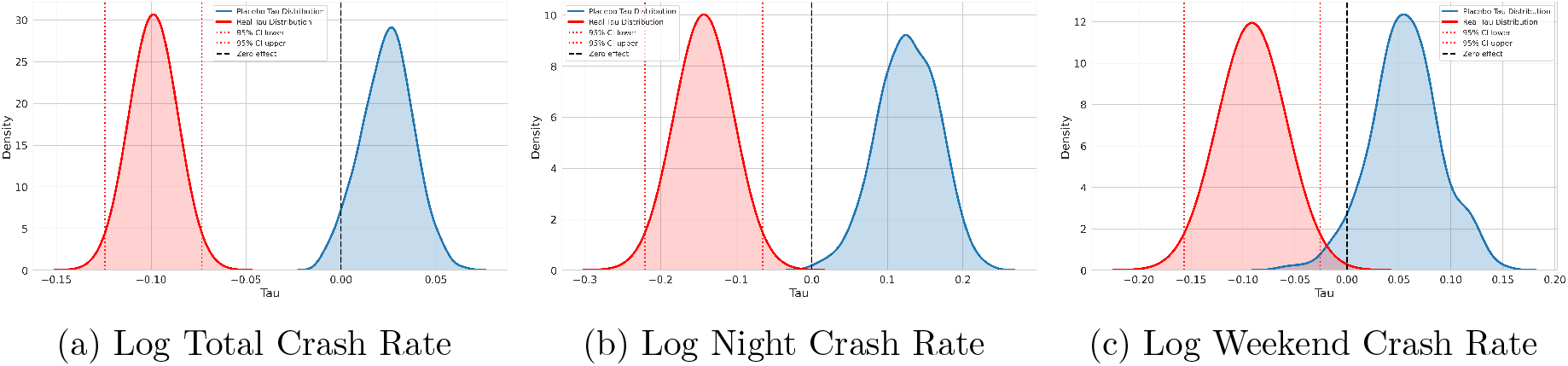
Real and placebo tau distributions in Cluster 0

**Figure 26:**
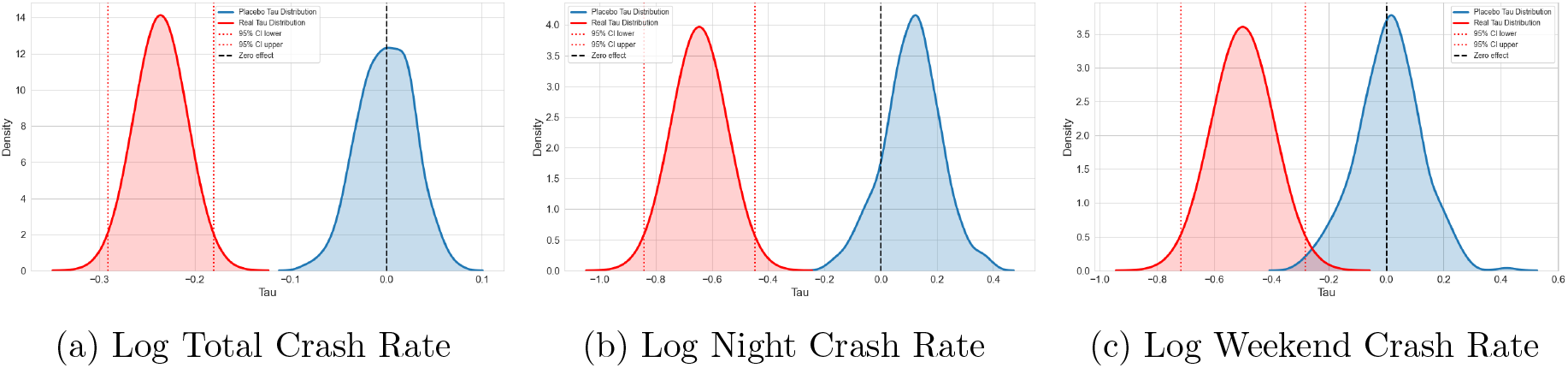
Real and placebo tau distributions in Cluster 1

**Figure 27:**
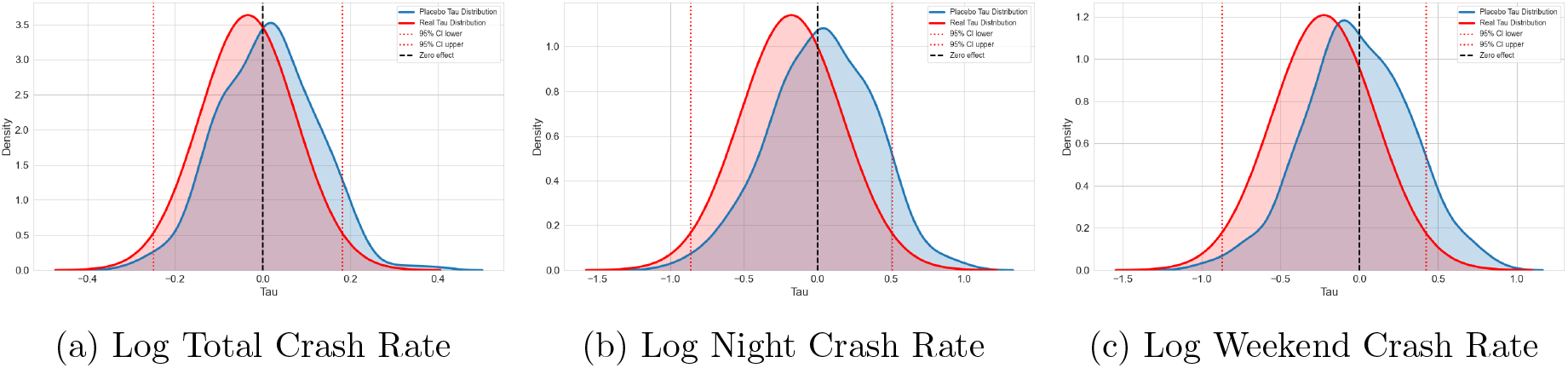
Real and placebo tau distributions in Cluster 2

**Figure 28:**
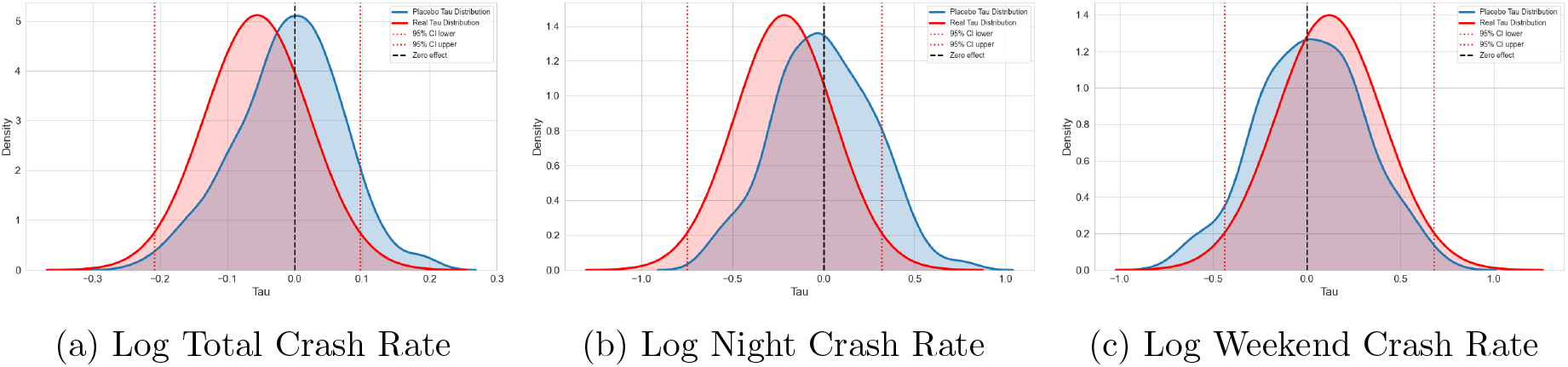
Real and placebo tau distributions in Cluster 3

**Figure 29:**
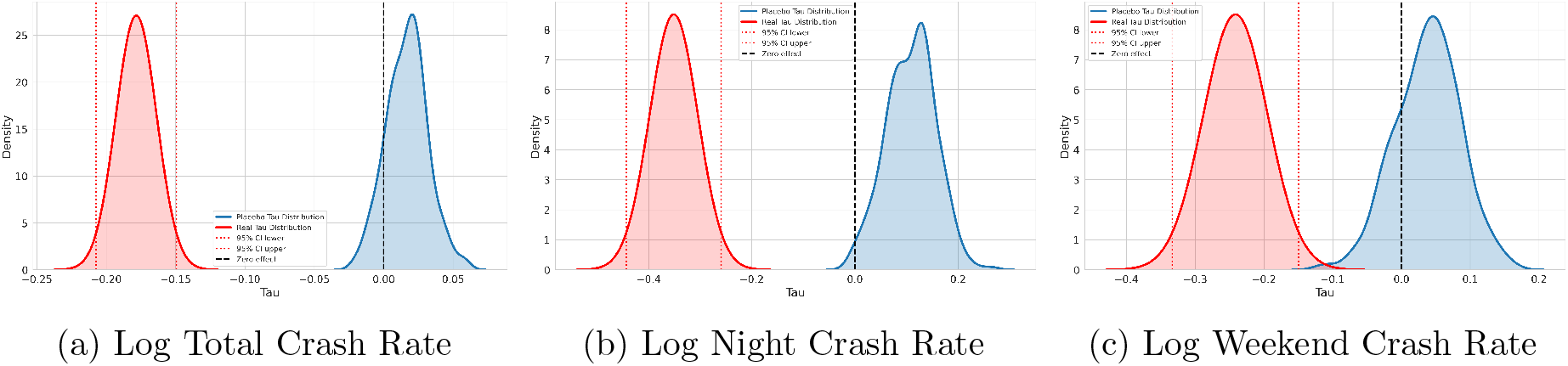
Real and placebo tau distributions in Cluster 4

### D.1 Aggregate Effects

### D.2 Heterogeneous Effects

## Footnotes

1 Available at https://tinyurl.com/bdcs8rnn.

2 Available at https://tinyurl.com/jspmdr63.

3 Available at https://tinyurl.com/4vyjsccc.

4 Available at https://tinyurl.com/ytuttxkp.

5 Available at https://tinyurl.com/42xnu88a.

6 Available at https://tinyurl.com/yyyv8nxe.

7 Available at https://tinyurl.com/yc5tkysr.

8 Available at https://tinyurl.com/y5593v8r for England and Wales.

9 Available at https://tinyurl.com/mr2k33hn for Scotland.

10 Available at https://tinyurl.com/2jyf5kam.

## Notes

### Competing Interest Statement

The authors have declared no competing interest.

### Author Declarations

This study uses anonymized, routinely collected administrative data obtained from publicly available sources (all the links are available in the manuscript). No identifiable individual-level information was accessed, and no participants were directly recruited or contacted. Ethical approval was therefore not required under institutional and national guidance.

